# Help-seeking needs related to suicide prevention for individuals in contact with mental health services: A rapid scoping review

**DOI:** 10.1101/2024.07.11.24310222

**Authors:** Hwayeon Danielle Shin, Jessica Kemp, Samantha Groves, Laura Bennett-Poynter, Charlotte Pape, Karen Lascelles, Gillian Strudwick

## Abstract

**Introduction:** Prior mental health care utilization presents an important window of opportunity for providing suicide prevention interventions. To date, no reviews have consolidated the help-seeking needs of individuals in contact with mental health services. This warrants further attention given this group may have different needs for interventions compared to the general population who have not sought help previously.

**Aim:** The purpose of this rapid scoping review was to summarize the available literature on help-seeking needs related to suicide prevention among individuals in contact with mental health services from healthcare settings.

**Method:** Cochrane rapid review and Joanna Briggs Institute scoping review methodologies were adapted, and databases, including MEDLINE, Scopus, CINAHL, PsycInfo, and EMBASE, were searched.

**Results:** A total of 42 primary studies were included in analysis. Reported barriers and facilitators to help-seeking behaviors identified within studies were mapped onto the socio-ecological model. Barriers and facilitators identified included knowledge and attitudes toward healthcare utilization, family and peer support, interactions with healthcare professionals, provision of holistic care, and the creation of a supportive atmosphere and safe space to promote open discussions of suicide-related concerns.

**Discussion:** The findings of this review offer valuable insights into areas for improvement in addressing help-seeking needs for individuals who are in contact with health services related to suicide prevention.

**Implication for Research:** The findings serve as a foundation for shaping mental health initiatives informing approaches and care delivery tailored towards individuals who are in contact with health services. The reported barriers and facilitators offer insights to inform the development of mental health support tools to enhance care and considerations for evaluations.

**Accessible Summary:** *What is known on the subject:*

- When individuals contact healthcare services while experiencing suicidal thoughts or behaviors, it is an important opportunity to offer them help and interventions to prevent suicide; however, this does not necessarily mean that their needs are always met.
- Previous research has identified abundant evidence on help-seeking barriers and facilitators for mental health support; however, we do not have a consolidated understanding of the help-seeking needs, including unmet needs, for individuals in contact with mental health services, which may differ from individuals who have not sought care recently.

*What the paper adds to existing knowledge:*

- This review consolidated the reasons why health service users might seek or avoid help when experiencing thoughts of suicide, organizing them as barriers and facilitators within the adapted socio-ecological model.

*What are the implications for research:*

- The findings from the review can form the basis for shaping mental health initiatives related to approaches and care delivery.
- The identified barriers and facilitators can provide valuable insights for designing mental health support tools and considerations for evaluations.

## Background

Every year, around 700,000 people die by suicide globally (World Health Organization, 2014, 2021). Based on an analysis of the World Mental Health Surveys (conducted 2001-2007) involving 108,705 adults in developed countries, the estimated 12-month prevalence of suicidal thoughts, plans, and attempts was 2.0%, 0.6%, and 0.3% respectively (Borges *et al*., 2010). A more recent meta-analysis of 66 studies including children and adolescents estimated the global 12-month prevalence of suicidal thoughts, plans and attempts to be 14.2%, 7.5% and 4.5% respectively (Lim *et al*., 2019). In line with these findings, evidence continues to highlight the significant prevalence of suicide-related thoughts and behaviours among adults and youth worldwide (Lovero *et al*., 2023; Van Meter *et al*., 2022), emphasizing the need for effective suicide prevention and mental health support strategies.

Previous research has identified an abundance of evidence on risk factors for suicide and suicidal behaviors (Favril *et al*., 2023). Previous self-harm or suicide attempts are the strongest risk factors for future suicide (Hawton *et al*., 2015, 2020), with mental health conditions also contributing to risk (Favril *et al*., 2022). A study reviewing 2,674 patients who died by suicide from eight healthcare systems found that 51.3% of individuals had at least one mental health diagnosis recorded in the year before their death. The diagnoses associated with highest risk of suicide were schizophrenia, bipolar, depressive, and anxiety disorders followed by attention deficit hyperactivity disorder (ADHD) (Yeh *et al*., 2019). Among individuals with mental health conditions or a history of previous suicidal behaviors, demographic characteristics may increase or decrease risk of suicide including, but not limited to, gender (Marconi *et al*., 2023; Miranda-Mendizabal *et al*., 2019), age (Biswas *et al*., 2020; Clements *et al*., 2019; Hawton *et al*., 2020; Murphy *et al*., 2012), sexual orientation (Liu *et al*., 2019), socio-economic status (Cairns *et al*., 2017; Qian *et al*., 2023), experiences of marginalization (Pollock *et al*., 2018; Rudes & Fantuzzi, 2022), geographic location (Knipe *et al*., 2022), neurodiversity and autism (Cassidy *et al*., 2020), physical health (Qin *et al*., 2022), and occupation (Windsor-Shellard & Gunnell, 2019).

Suicide is preventable through evidence-based interventions (Mann *et al*., 2005; Nuij *et al*., 2021; Zalsman *et al*., 2016). Connecting individuals experiencing suicidal ideation and behaviors with appropriate mental health services is a critical aspect of suicide prevention and support efforts. Many individuals who die by suicide or attempt suicide are already in contact or have been in contact with mental health services (Stene-Larsen & Reneflot, 2019). A systematic review examining rates of contact with primary and mental health services prior to suicide found that among individuals who died by suicide, 57% had received mental health care in their lifetime, and 31% had received care in the year before death (Stene-Larsen & Reneflot, 2019). This finding is consistent with studies in other countries such as Wales (John *et al*., 2020) and Sweden (Bergqvist *et al*., 2022). The recency of healthcare utilization prior to suicide presents a key window of opportunity for individuals to be provided with suicide prevention interventions and support.

## Rationale and aim

Previous reviews have examined help-seeking behaviours regarding mental health among individuals who are at increased risk of suicidal behaviours. A systematic review examining help-seeking behaviors of adolescents for common mental health issues (including suicidal ideation) found stigma and unfavorable perceptions of mental health services were primary barriers to seeking help (Aguirre Velasco *et al*., 2020). In contrast, prior positive experiences with health services and increased mental health awareness were help-seeking facilitators (Aguirre Velasco *et al*., 2020). Another review focusing on adults looked at help-seeking behaviors related to mental health needs (Adams *et al*., 2022). The most frequently reported influencing factors included patients’ attitudes and perceived behavioral control, both of which were significant predictors of help-seeking intentions (Adams *et al*., 2022). Other studies have identified additional factors which may play a significant role in influencing help-seeking behaviors. These include knowledge, financial resources, gender or occupation-specific stereotypes, and structural barriers/facilitators, such as place of residence (Gulliver *et al*., 2010; Radez *et al*., 2021; Randles & Finnegan, 2022). However, what remains unclear is the consolidated understanding of the help-seeking behaviours and barriers and facilitators to help-seeking among individuals who are in contact with mental health services. This is important to know since there may be unique interventions or supports available within the context of service delivery, which may differ from those aimed at individuals who have not sought care previously.

The overall aim of this rapid scoping review is to describe the barriers and facilitators to help-seeking for suicide prevention care among individuals who are in contact with and/or accessing mental health services in healthcare settings. Understanding these are essential for improving quality of care, reducing suicide risk, and ensuring that patient needs are met – all of which contribute to the ongoing improvement of mental health services. Furthermore, this review is part of a broader project aiming to inform a new iteration of a digital tool, which will be designed to support suicide prevention care for individuals in contact with mental health services. Before developing the new iteration of the tool, our goal is to identify currently unaddressed help-seeking needs among individuals who are in contact with mental health services.

A preliminary search of MEDLINE was conducted in March 2023, which did not identify any current or in-progress scoping or systematic reviews on this topic.

### Review questions

1. What are the characteristics of studies that investigate help-seeking needs related to suicide prevention care for individuals who are in contact with mental health services?
2. What are the barriers and facilitators of help-seeking related to suicide prevention care among individuals in contact with mental health services?

## Methods

We conducted a rapid scoping review, adapting the standard methodological guidelines for rapid (i.e., Cochrane Rapid Reviews Methods Group guidelines (Garritty *et al*., 2021)) and scoping reviews (i.e., Joanna Briggs Institute (Peters *et al*., 2015, 2020)). This type of review can be beneficial to inform urgently needed practice. In this case, the review was conducted to urgently inform a new iteration of a digital tool to support suicide prevention care in mental health services. Researchers employ various approaches to conduct rapid reviews without compromising rigor, aiming to streamline the review process to meet the urgent needs (Tricco *et al*., 2015). Here, we expedited the process by narrowing our search scope to the most relevant databases for the topic, establishing strict eligibility criteria, involving two independent reviewers in titles, abstracts, and full-text screening processes, and excluding the assessment of study quality. There was no a priori protocol registered, and we prepared this review report adhering to the Preferred Reporting Items for Systematic Reviews and Meta-Analyses extension for Scoping Reviews guidelines (PRISMA-ScR) (Tricco *et al*., 2018).

### Eligibility criteria

#### Concept

The primary concepts under investigation were suicide, suicidal ideation/thoughts, and suicide attempts. We followed the international definitions suggested by De Leo et al. (2021) to define the above suicide phenomena (De Leo *et al*., 2021). Studies examining non-suicidal self-injury (i.e., self-harm behaviours and injuries without suicide intent) were ineligible for inclusion. However, in some studies, suicide attempts were identified by self-harm codes (e.g., in emergency department records). To be included, studies were required to have at least one of these items as an outcome of interest. Additionally, studies were expected to describe help-seeking needs, barriers or facilitators to help-seeking, or unmet needs related to suicide prevention. Help-seeking was defined as a planned behaviour of actively seeking help from a healthcare professional (or crisis line worker) initiated by recognizing an issue or changes in health, in this case relating to mental health (Cornally & McCarthy, 2011). Help-seeking for mental health is influenced by a combination of social, individual, and structural factors, as mental health concerns are intricately tied to their surroundings and seeking help is navigated through social engagement and cultural practices (Adams *et al*., 2022; Jang *et al*., 2007; Martinez *et al*., 2020; Mojtabai, 2009).

#### Population and Context

Herein, we define mental health services contact as either a visit or a phone call to any mental health care professionals in healthcare settings. This typically indicates that individuals are actively engaged with or seeking assistance from healthcare providers or institutions, and it can also mean accessing services and receiving treatment. To be eligible for inclusion, studies needed to focus on individuals in contact with mental health services from healthcare settings, which could include in-patient, out-patient, primary care settings, emergency department, and crisis line services. We included emergency departments because individuals seeking mental health services can access care through the emergency department, where they undergo assessments by healthcare providers for evaluation, stabilization, counseling, medication, or referral to specialized mental health services. Additionally, we included crisis lines because they offer a way to connect individuals in distress with trained responders, with a special focus on suicide prevention. However, it is important to acknowledge that contact with health services does not always result in the receipt of treatment, highlighting unmet help-seeking needs.

Additionally, our primary interest was exploring help-seeking needs subsequent to contact with mental health services. When the timing of this contact was unclear, we determined temporality based on information provided by the authors. Studies were included if individuals were in contact with healthcare services either before or at the time of the study. However, studies focusing solely on individuals managing suicide-related thoughts and behaviors independently, without utilizing any formal healthcare services and relying solely on self-help tools or informal carers (e.g., family or friends), were excluded.

#### Search Strategy

Our search strategy was developed with a health science librarian and followed the PRESS guidelines (McGowan *et al*., 2016). Before full searches were conducted, an initial limited search of MEDLINE was undertaken to identify relevant articles, and then text contained in the titles and abstracts alongside index terms of identified articles were used to inform development of the final search strategy. The search strategy, including keywords and index terms, was adapted for the following databases: MEDLINE, Scopus, CINAHL, PsycInfo, and EMBASE. Searches were conducted on 9^th^ May 2023, and the full search strategies are provided in Supplementary File 1.

### Study selection

All records identified by database searching were collated and uploaded into Covidence, a software to support literature screening (“Covidence systematic review software,” 2019), and system-detected duplicates were automatically removed. Following piloting to ensure consistency in screening, titles and abstracts were screened by two independent reviewers (HDS, JK, SG, LBP, CP) against the inclusion criteria. Potentially relevant articles were retrieved in full, and two independent reviewers (HDS, JK, SG, LBP, CP) read each full-text articles for detailed inclusion assessment. Ineligible articles were excluded, and reasons for their exclusion were documented. Any interpretation disagreements that arose between the reviewers were resolved through team discussion.

### Data extraction

Data were extracted from included articles by two of the five reviewers (HDS, JK, SG, LBP, CP) using the data extraction tool on Covidence. Two independent reviewers initially piloted the tool (HDS, JK) using two articles to assess the correctness and completeness of the extracted data. After ensuring the consistency of data extraction, remaining articles were extracted by a single reviewer, and validated by a second reviewer. General characteristics of the article (e.g., author, publication country), design and methods including data sources, participant characteristics including considerations for intersectionality to report the prevalence of inequalities, outcome of interest, and barriers and facilitators for help-seeking, were extracted. See Supplementary File 2 for a full list of items in the data extraction tool. Any interpretation disagreements that arose during the validation step were resolved through discussion between reviewers.

### Data analysis and presentation

Data analysis was carried out independently by a single reviewer (HDS, JK, SG, LBP, CP) and validated by a second reviewer. We used a directed content analysis approach (Hsieh & Shannon, 2005) to characterize extracted narrative data. Data is presented narratively and in table format.

During analysis, participant characteristics were extracted and organized using the intersectionality framework (Crenshaw, 1989, 1991). The framework recognizes how social identities are interconnected and can lead to experiences of different levels of social power and positions, encompassing both privilege and oppression. As some groups, such as middle-aged men (Qin *et al*., 2022), sexual minority groups (Kidd *et al*., 2023), adolescents (Biswas *et al*., 2020), and individuals with mental health conditions (Yeh *et al*., 2019), have high prevalence of suicidal behaviours, we extracted data on these intersecting identities to gather a comprehensive understanding of unmet needs among subgroups of individuals in contact with mental health services. During the extraction process, we recognized that some authors interchangeably used the terms, ‘gender’ and ‘sex’; for example, authors used women/men or female/male to define both gender and sex. To facilitate accurate reporting, we exercised our best judgment based on the categories as reported by the authors.

Extracted barriers and facilitators to help-seeking were organized using the adapted ecological model (McLeroy *et al*., 1988), categorizing them into the following five distinct, yet not mutually exclusive levels: 1) Individual; 2) Interpersonal; 3) Organizational/Institutional; 4) Community and Societal; and 5) National Policy/Law. Individual factors encompassed participant characteristics such as knowledge, behaviors, skills, judgement, and attitudes including perceived opinions. Interpersonal factors encompassed interactive processes and relationships within primary social groups, whether formal or informal social networks, and support systems including families, workgroups, and friendship networks. Institutional and organizational factors encompassed characteristics of institutions and formal/informal operational rules and regulations, such as training support for clinicians in hospitals. Community and societal factors encompassed relationships among organizations, institutions, wider social networks and informal networks within defined boundaries. National, policy and law levels encompassed local, state, or national laws and policies, for example, health insurance concerns. It was necessary for reviewers to interpret information to a certain extent, relying on what the authors had reported. Differences in interpretation identified during the validation process were addressed through discussion between reviewers.

## Results

### Study inclusion

Our search strategy yielded 2,493 citations (Figure 1). After automatic removal of duplicates, 1,755 articles were screened based on title and abstract. Of these, 128 articles proceeded to full-text evaluation, ultimately resulting in 42 eligible studies. Reasons for exclusion were documented and they are outlined in the PRISMA flow chart (Figure 1).

**Figure 1.**
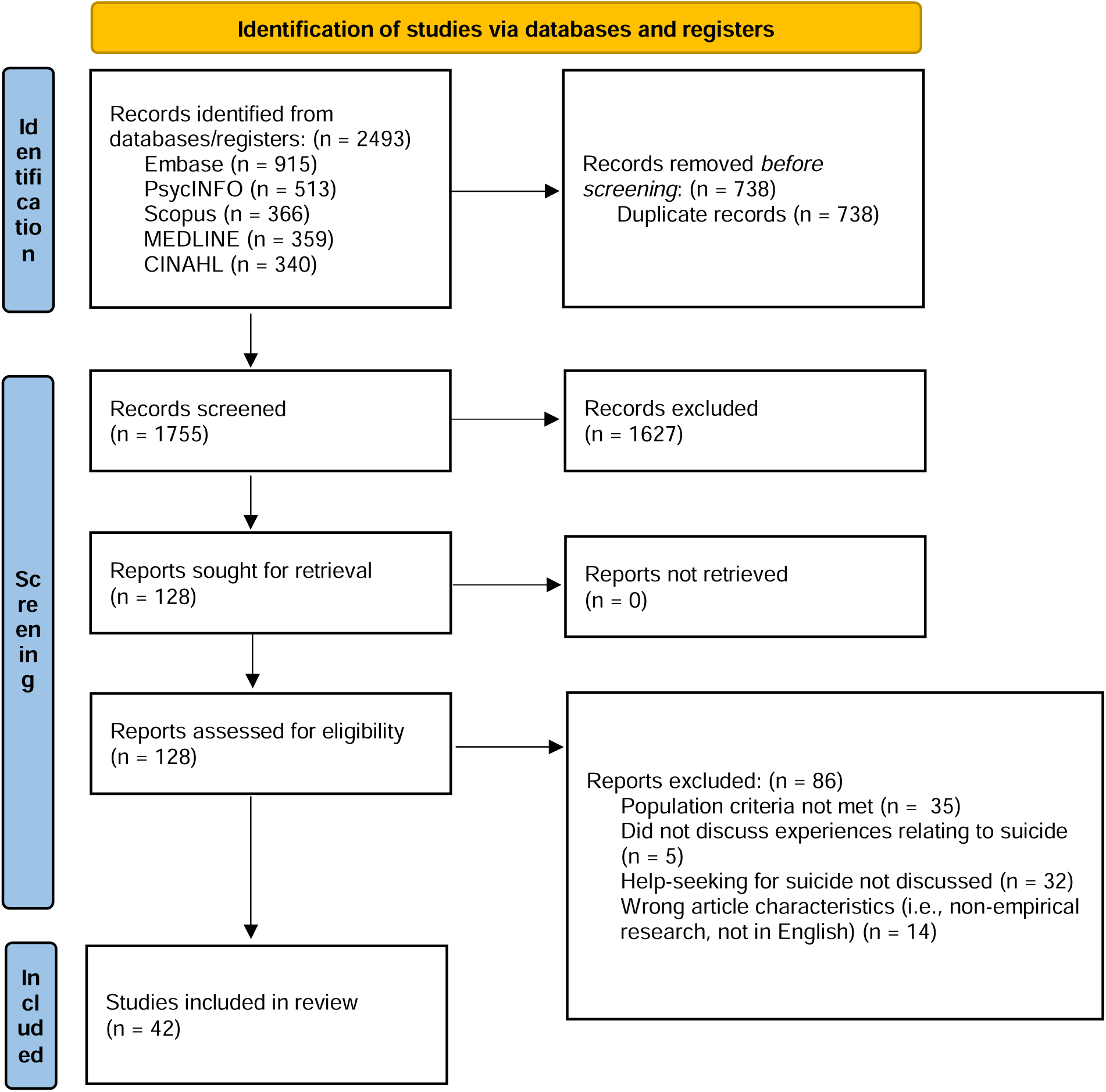
PRISMA flow chart

### Characteristics of included studies

Individual characteristics of included citations are organized in Table 1. The majority of studies were observational (n=19) and qualitative (n=17), followed by mixed/multi-methods (n=3), experimental (n=2), and quasi-experimental (n=1). Nearly half (n=19) of included studies were published between 2011 to 2020, and 13 were published within the last three years (2021-2023). Studies originated from the United States (n=16), United Kingdom (n=6), Australia (n=4), Canada (n=3), Sweden (n=3), Norway (n=2), Netherlands (n=2) and elsewhere in Asia, Europe, East Africa, and Oceania (see Table 2 for a summary of included studies). Suicide phenomena examined included suicide attempt (i.e., behaviour) (n=34), suicide ideation (n=26), and death by suicide (n=7). Data sources included interviews (n=23), surveys (n=14), health administrative data (n=8), and other sources (n=5), such as data obtained from text-based or call-line crisis services. Included studies commonly reported on demographic characteristics of participants including age (n=35), sex (n=28), marital status (n=17), gender (n=15), race (n=13), ethnicity (n=11), and income (n=10). Eighteen studies reported other characteristics such as previous mental health diagnosis, medical history, and employment status. See Figure 2 for a full list of characteristics reported in included studies.

**Figure 2.**
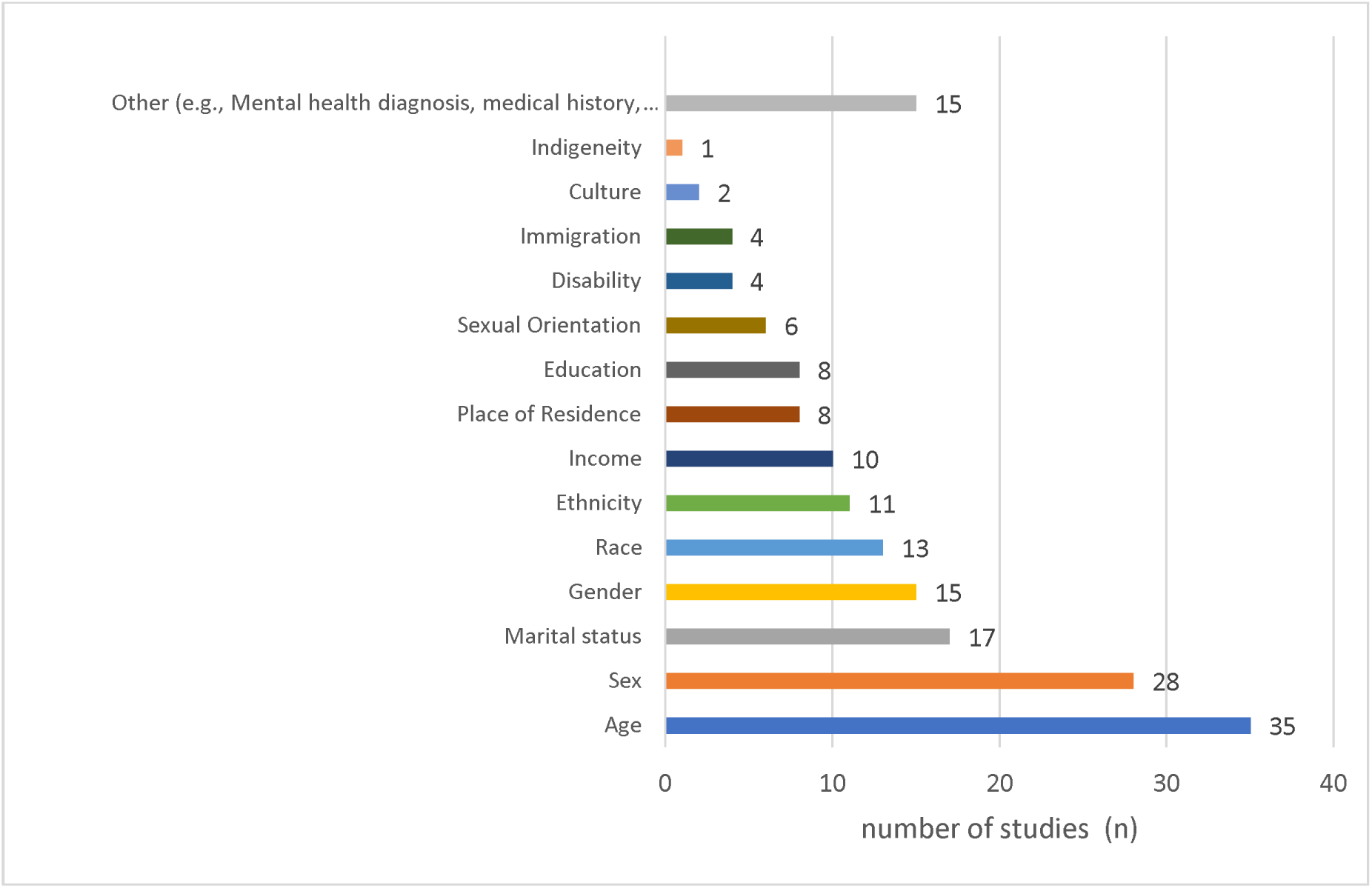
Reported characteristics of participants

**Table 1.** Study characteristics.

| Author, Year | Country of Origin | Design | Aim of Study | Sample Description | Suicide Phenomena | Health Services Setting | Data Source |
| --- | --- | --- | --- | --- | --- | --- | --- |
| (Adler *et al.*, 2020) | USA | Qualitative | Examine barriers to treatment among Army soldiers who experienced a recent suicidal crisis and had a history of deployment | 12 army soldiers receiving psychiatric inpatient treatment | Suicide attempt/behaviour;<br>Suicidal ideation | Inpatient psychiatric unit | Interview |
| (Amin *et al.*, 2021) | Sweden | Observational | Investigate if the patterns of inpatient and specialised outpatient psychiatric and somatic healthcare use differ between refugees and Swedish born individuals 3 years before and after a suicide attempt<br>Examine whether healthcare use differs among refugees as a result of sex, age, education or receipt of disability pension | 85,771 individuals who had attempted suicide and were treated by specialist healthcare<br>(81,916 Swedish-born, 3,855 refugees) | Suicide attempt/behaviour | Specialist healthcare services (inpatient and outpatient) | Health admin |
| (Ammerman *et al.*, 2021) | USA | Observational | Explore how experiences and reactions related to disclosing a prior suicide attempt relates to future help-seeking outcomes | 37 veterans with previous suicide attempt receiving psychiatric inpatient treatment | Suicide attempt/behaviour | Inpatient psychiatric unit | Survey |
| (Barnes *et al.*, 2002) | USA | Observational | Describe the help-seeking behaviors engaged in by adolescents and young adults prior to a nearly lethal suicide attempt | 153 adolescents and young adults (aged 13-34) treated at an emergency hospital following a suicide | Suicide attempt/behaviour | Emergency department | Survey |
|  |  |  | Examine the association between help-seeking behaviors and nearly lethal suicide attempts | attempt |  |  |  |
| (Barry *et al.*, 2023) | Canada | Observational | Determine the number of individuals who seek help prior to suicide or a suicide attempt, separated by urban or rural status<br>Examine the association between rurality and help-seeking among individuals who have died or attempted suicide | 9,848 individuals who died by suicide identified through emergency department data and verified by coronial data.<br>82,480 individuals who attempted suicide and presented to an emergency department | Death by suicide; Suicide attempt/behaviour | Emergency department | Health admin (coronial data for death by suicide) |
| (Camgan, 1994) | UK (Northern Ireland) | Qualitative | Investigate the psychosocial needs, of individuals who have attempted suicide by self-poisoning<br>Evaluate healthcare support received during and following the suicide attempt | 6 individuals who were in hospital following a suicide attempt by intentional overdose | Suicide attempt/behaviour | Emergency department | Interview |
| (Cedereke & Öjehagen, 2007) | Sweden | Qualitative | Examine formal and informal help related to clinical and social needs that patients who attempted suicide received the<br>year following their suicide attempt<br>Examine whether help from services was perceived to be adequate by patients<br>Investigate whether patients who had repeat suicide attempts during the follow-up differed from those who did not | 140 individuals who had been treated at a medical emergency inpatient unit following a suicide attempt | Suicide attempt/behaviour | Medical emergency inpatient unit | Interview |
| (Chia-Yi Wu *et al.*, 2012) | Taiwan | Qualitative | Explore how and why people with deliberate self-harm access formal or informal help sources and related factors | 20 individuals who presented to the emergency department following self-harm and were referred to a suicide prevention center | Suicide attempt/behaviour | Suicide prevention centre following emergency department presentation | Interview |
| (Chung, 2010) | USA | Qualitative | Examine the interplay of informal and formal help-seeking processes within a sample of Chinese immigrants in the USA | 31 Chinese immigrants who enrolled for treatment at a mental health clinic following a suicide attempt after re-settling in USA | Suicide attempt/behaviour | Mental health clinics | Interview |
| (Cleary, 2017) | Ireland | Qualitative | Examine psychiatric service use and attitudes to treatment among a sample of men who made a medically serious | 52 men who presented to three hospitals in Ireland following a medically serious | Suicide attempt/behaviour | Emergency department, inpatient psychiatric unit, | Interview |
|  |  |  | suicide attempt. | suicide attempt. |  | outpatient services |  |
| (de Lange *et al.*, 2022) | Netherlands | Qualitative | Explore experiences and needs related to formal and informal mental health care for sexual and gender minority youth who were experiencing suicidal ideation. | 23 young adults who identified as sexual gender minority group (aged 18-35) | Suicide attempt/behaviour;<br>Suicidal ideation | Most young adults interviewed received mental health care from a healthcare professional (e.g., psychologist, psychiatrist) | Interview; Focus group |
| (Denneson *et al.*, 2010) | USA | Observational | Describe health care contacts at a Department of Veterans Affairs medical center in Oregon in the year prior to the death of veterans by suicide | 112 veterans who died by suicide in Oregon and had contact with a single Veterans Affairs Medical Centre in the year before their death | Death by suicide;<br>Suicide attempt/behaviour;<br>Suicidal ideation | Veterans Affairs Medical Center | Health admin |
| (Hagen *et al.*, 2020) | Norway | Mixed Methods | Explore how patients experience suicidality and their experience of being in a psychiatric hospital | 11 women who were admitted to a psychiatric hospital and were experiencing suicidality | Suicide attempt/behaviour;<br>Suicidal ideation | Psychiatric acute section | Interview;<br>Other:<br>Participant observation in the hospital. |
| (Hausmann-Stabile *et al.*, 2018) | USA | Mixed methods (Qualitative Report) | Examine the pathways related to and characteristics of services provided to adolescents following suicidal crisis, and examine what the adolescent's experiences of care | 68 Latina adolescents (aged 11-19) who had attempted suicide and had contact with medical and mental health services | Suicide attempt/behaviour;<br>Suicidal ideation | Range of mental health services (e.g., inpatient, outpatient) | Interview |
| (Heinsch *et al.*, 2020) | Australia | Qualitative | Explore the experience of individuals with comorbid depression and alcohol use disorders who had previously attempted suicide regarding the help they sought and received prior to and following the attempt | 6 individuals with a prior suicide attempt who had participated in the Self-Help for Alcohol and other drug and Depression (SHADE) study | Suicide attempt/behaviour;<br>Suicidal ideation | Range of mental health services (e.g., mental health professional, phone helpline) | Interview |
| (Holt *et al.*, 2023) | USA | Qualitative | Explore the lived experience of sexual and gender minority individuals who have had a recent near fatal suicide attempt. Specific focus on help-seeking and previous mental health care experiences | 22 individuals who identified as sexual and gender minority group and had reported at least one near-fatal suicide attempt in the past 18 months. | Suicide attempt/behaviour;<br>Suicidal ideation | Range of mental health services (e.g., inpatient, outpatient, helpline) | Interview |
| (Hom *et al.*, 2018) | USA | Observational | Identify the rates of professional and nonprofessional mental health service use and help-seeking, alongside preferred sources of support among female firefighters with a history of suicidality<br>Identify possible barriers to treatment seeking for psychological problems and correlates of mental health service use among this group | 119 female firefighters who have a history of suicidal thoughts, plans, or attempts | Suicide attempt/behaviour;<br>Suicidal ideation | Range of mental health services (e.g., psychologist, therapist, counselor, psychiatrist, and other medical providers including physicians and nurse practitioners) | Survey |
| (Ilgen *et al.*, 2021) | USA | Experimental | Analyze the immediate effect of the Crisis Line Facilitation intervention on self-reported help seeking for suicidal crisis among veterans. | 301 veterans receiving psychiatric inpatient treatment for a recent suicidal crisis (defined as significant ideation, plan, and/or recent attempt requiring psychiatric hospitalization) | Suicide attempt/behaviour;<br>Suicidal ideation | Crisis line and inpatient psychiatric care | Survey;<br>Interview |
| (John *et al.*, 2020) | UK (Wales) | Observational | Examine the type and timing of healthcare contacts in individuals who died by suicide in Wales | 5130 individuals who died by suicide identified using The Suicide Information Database-Wales<br>1721 suicide deaths were eligible for full-linked analysis | Death by suicide | Range of health services (e.g., general practice, emergency department, secondary care) | Health admin |
| (Jones *et al.*, 2021) | USA | Qualitative | Explore the impact of involuntary hospitalization on youth and young adult's trust, help-seeking and engagement with clinicians | 40 youth and young adults (aged 16-27) who had been involuntarily hospitalized to a psychiatric hospital. | Suicide attempt/behaviour;<br>Suicidal ideation | Psychiatric hospital | Interview |
| (Jordan *et al.*, 2012) | UK (Northern Ireland) | Qualitative | Determine the preferences, and key features and processes of appropriate services and clinical care for young men who are suicidal | 36 formerly suicidal young men | Suicide attempt/behaviour;<br>Suicidal ideation | Range of mental health services (e.g., inpatient, outpatient,<br>community mental<br>health nursing,<br>counseling) | Interview |
| (Jordans *et al.*, 2018) | Netherlands | Observational | Identify the prevalence of non-fatal suicidal behaviour within community and health facility based populations in five low- and middle-income countries | 6470 individuals presenting at primary care facilities | Suicide attempt/behaviour;<br>Suicidal ideation | Primary care | Survey |
| (Karras *et al.*, 2017) | USA | Experimental (Pilot) | Evaluate messaging strategies to enhance use of the Veterans Crisis Line | The overall Veterans Crisis Line call rate for all pilot city sites was 4.81 per 1,000 veterans (exact total call number not provided). | Suicide attempt/behaviour;<br>Suicidal ideation | Crisis line | Other: Veteran Crisis Line call data. |
| (Kodama *et al.*, 2016) | Japan | Quasi-experimental | Assess a mobile phone intervention, with a focus on help-seeking and self-harming behaviours | 30 psychiatric outpatients who had experienced suicidal ideation | Suicide attempt/behaviour;<br>Suicidal ideation | Outpatient psychiatric care | Health admin;<br>Survey |
| (Kuramoto-Crawford *et al.*, 2017) | USA | Observational | Examine self-reported reasons for not receiving mental health treatment among adults with suicidal thoughts in the past year<br>Examine whether sociodemographic characteristics and health insurance status was associated with reasons | 8400 American citizens completing the National Surveys on Drug Use and Health Experiencing suicidal ideation or previous attempt | Suicide attempt/behaviour;<br>Suicidal ideation | Individuals considered seeking help but could not gain access due to various self-reported reasons | Survey |
| (Leavey *et al.*, 2016) | UK<br>(Northern Ireland) | Observational | Examine predictors of health and social care service contact in the 12-month period prior to suicide and General Practitioner detection of suicidality | 361 Suicide deaths recorded by the Coroner's Office | Death by suicide | General Practitioner services and a range of mental health services (e.g., nursing services, social workers, emergency services) | Health admin |
| (Lueck & Poe, 2021) | USA | Observational | Identify psychological determinants of utilizing a helpline and examine potential barriers in order to inform effective help-line promotion | 22 Students (currently in counseling) | Death by suicide | Helpline | Survey |
| (McKay & Shand, 2018) | UK | Qualitative | Examine the stories of adults who have previously attempted suicide including exploring experiences of Australian healthcare services | 20 individuals who had attempted suicide in the previous 18 months | Suicide attempt/behaviour | Following suicide attempt, all but one individual received mental health care. | Interview |
| (Pavulans *et al.*, 2012) | Sweden | Qualitative | Explore the lived experience of being suicidal and having made a suicide attempt to identify potential implications for health care professionals concerning how to meet, care for and treat people who have made a suicide attempt and help them not to attempt suicide again | 10 individuals who had recently attempted suicide (within 3 weeks) and were receiving psychiatric inpatient treatment | Suicide attempt/behaviour | Inpatient psychiatric unit | Interview |
| (Pirkis *et al.*, 2001) | Australia | Observational | Examine the self-reported needs of users of mental health services who are suicidal (and non-suicidal), and examine whether these needs of met | 245 individuals with suicidal ideation (from a total survey pool of 10,641 adults). | Suicide attempt/behaviour; Suicidal ideation | Range of mental health services | Survey |
| (Podlogar *et al.*, 2022) | USA | Observational | Investigate whether certain level of past mental health care receipt is associated with later explicit nondisclosure of suicide risk to a health care professional<br>Explore whether severity of current suicidal ideation and other hypothesized psychological constructs (i.e., willingness to seek help for suicidal thoughts) may impact any associations | 282 men aged 50+ with varied history of previous mental health care utilisation | Suicide attempt/behaviour; Suicidal ideation | Inpatient and outpatient mental health care | Survey |
| (Rainbow *et al.*, 2023) | Australia | Observational | Evaluate factor structure, internal consistency, convergent and divergent validity of Suicide-Related Coping Scale (SRCS) in two Australian online help-seeking samples | 1,266 visitors to a mental health website who are experiencing suicidal ideation<br>693 users of a suicide safety planning mobile app | Suicidal ideation | Users of a suicide prevention mobile app, and visitors to a mental health website | Survey |
| (Raingruber, 2002) | USA | Qualitative | Explore issues that influence the care of depressed and suicidal individuals from the perspective of clients, licensed mental health professionals, primary care physicians, police officers, high school teachers, counselors, and parents | 10 mental health care clients | Suicide attempt/behaviour;<br>Suicidal ideation | Range of mental health services | Interview |
| (Renaud *et al.*, 2014) | Canada | Mixed methods | Determine what health care services were received by individuals who died by suicide, and what healthcare needs these individuals had | 67 individuals who had died by suicide who were aged 25 or younger | Death by suicide | Range of health services (e.g., medical services, allied health, specialized mental health services, helpline, etc.) | Health admin;<br>Interview |
| (Rheinberger *et al.*, 2021) | Australia | Qualitative | Understand the experiences of help seekers when presenting to the emergency department as a result of suicidal crisis. A focus on what works and doesn't work, and areas for improvement of care | 17 individuals who had presented to a emergency department as a result of suicidal crises within the past 18 months | Suicide attempt/behaviour;<br>Suicidal ideation | Emergency department | Interview |
| (Saini *et al.*, 2021) | UK (England) | Observational | Compare help-seeking among younger and older men who attended a therapeutic centre for men who are in suicidal crisis | 546 Men attending a centre for men experiencing suicidal crisis. | Suicide attempt/behaviour | Suicide prevention centre | Health admin and survey;<br>Other: Questionnaires completed with therapist during session |
| (Skogstad *et al.*, 2006) | New Zealand | Observational | Assess whether prisoners' help seeking intentions related to a personal-emotional problem, such as suicidal feelings, can be predicted using variables from the Theory of Planned Behaviour | 527 male prison inmates | Suicidal ideation | Prison | Survey |
| (Spence *et al.*, 2008) | Canada | Qualitative | Investigate the experiences including needs and barriers to care alongside substance use history of men who have repeat presentations to emergency departments following suicidal crises | 25 males with suicide ideation or history of suicide intent with additional history of substance use problems, who had either been discharged from the emergency department, or admitted for inpatient psychiatric care | Suicide attempt/behaviour;<br>Suicidal ideation | Emergency department | Interview |
| (Talseth *et al.*, 1999) | Norway | Qualitative | Explore the experiences of individuals with prior suicide attempts or ideation related to their lived experience of care receipt from mental health nurses | 21 individuals receiving inpatient psychiatric care with previous suicide ideation or suicide attempt | Suicide attempt/behaviour;<br>Suicidal ideation | Psychiatric inpatient hospital | Interview |
| (Thompson *et al.*, 2018) | USA | Observational | Examine help-seeking behaviors by adolescent and young adults for a national text-based crisis support service | Adolescent crisis line users. 849,483 crisis Text Line conversation records | Death by suicide;<br>Suicidal ideation | Crisis text and phone line | Other: Text-based crisis service data |
| (Umubyeyi *et al.*, 2016) | Rwanda | Observational | Investigate help-seeking behaviours, barriers to care, and self-efficacy for help- | 247 young adults (aged 20-35) who were experiencing | Suicidal ideation | Range of health services (e.g., health centre or a | Interview |
|  |  |  | seeking among young adults with current depression and/or suicidality in a low-income setting | depression and/or suicidality |  | district hospital, trained mental health professional) |  |
| (Voros *et al.*, 2022) | Hungary | Observational | Assess the internet use patterns among psychiatric inpatients, with a focus on suicide-related content and its impact on help-seeking | 113 psychiatric inpatients with depressive disorders | Suicide attempt/behaviour; Suicidal ideation | Inpatient psychiatric unit | Survey |

**Table 2.** Summary of included studies.

| Characteristic | N | % | Citations of studies containing characteristic |
| --- | --- | --- | --- |
| **Study Design** |  |  |  |
| *Observational* | 19 | 45.2 | (Amin *et al.*, 2021; Ammerman *et al.*, 2021; Barnes *et al.*, 2002; Barry *et al.*, 2023; Denneson *et al.*, 2010; Hom *et al.*, 2018; John *et al.*, 2020; Jordans *et al.*, 2018; Kuramoto-Crawford *et al.*, 2017; Leavey *et al.*, 2016; Lueck & Poe, 2021; Pirkis *et al.*, 2001; Podlogar *et al.*, 2022; Rainbow *et al.*, 2023; Saini *et al.*, 2021; Skogstad *et al.*, 2006; Thompson *et al.*, 2018; Umubyeyi *et al.*, 2016; Voros *et al.*, 2022) |
| *Qualitative* | 17 | 40.4 | (Adler *et al.*, 2020; Camgan, 1994; Cedereke & Öjehagen, 2007; Chia-Yi Wu *et al.*, 2012; Chung, 2010; Cleary, 2017; de Lange *et al.*, 2022; Heinsch *et al.*, 2020; Holt *et al.*, 2023; Jones *et al.*, 2021; Jordan *et al.*, 2012; McKay & Shand, 2018; Pavulans *et al.*, 2012; Raingruber, 2002; Rheinberger *et al.*, 2021; Spence *et al.*, 2008; Talseth *et al.*, 1999) |
| *Mixed or Multi-Method* | 3 | 7.1 | (Hagen *et al.*, 2020; Hausmann-Stabile *et al.*, 2018; Renaud *et al.*, 2014) |
| *Experimental* | 2 | 4.8 | (Ilgen *et al.*, 2021; Karras *et al.*, 2017) |
| *Quasi- experimental* | 1 | 2.4 | (Kodama *et al.*, 2016) |
| **Country of Origin** |  |  |  |
| United States (US) | 16 | 38.1 | (Adler *et al.*, 2020; Ammerman *et al.*, 2021; Barnes *et al.*, 2002; Chung, 2010; Denneson *et al.*, 2010; Hausmann-Stabile *et al.*, 2018; Holt *et al.*, 2023; Hom *et al.*, 2018; Ilgen *et al.*, 2021; Jones *et al.*, 2021; Karras *et al.*, 2017; Kuramoto-Crawford *et al.*, 2017; Lueck & Poe, 2021; Podlogar *et al.*, 2022; Raingruber, 2002; Thompson *et al.*, 2018) |
| *United Kingdom*<br>*(including studies set in*<br>*individual UK countries*<br>*and across the UK)* | 6 | 14.3 | (Camgan, 1994; John *et al.*, 2020; Jordan *et al.*, 2012; Leavey *et al.*, 2016; McKay & Shand, 2018; Saini *et al.*, 2021) |
| Australia | 4 | 9.5 | (Heinsch *et al.*, 2020; Pirkis *et al.*, 2001; Rainbow *et al.*, 2023; Rheinberger *et al.*, 2021) |
| Canada | 3 | 7.1 | (Barry *et al.*, 2023; Renaud *et al.*, 2014; Spence *et al.*, 2008) |
| Sweden | 3 | 7.1 | (Amin *et al.*, 2021; Cedereke & Öjehagen, 2007; Pavulans *et al.*, 2012) |
| Norway | 2 | 4.8 | (Hagen *et al.*, 2020; Talseth *et al.*, 1999) |
| Netherlands | 2 | 4.8 | (de Lange *et al.*, 2022; Jordans *et al.*, 2018) |
| Japan | 1 | 2.4 | (Kodama *et al.*, 2016) |
| Ireland | 1 | 2.4 | (Cleary, 2017) |
| Taiwan | 1 | 2.4 | (Chia-Yi Wu *et al.*, 2012) |
| Rwanda | 1 | 2.4 | (Umubyeyi *et al.*, 2016) |
| New Zealand | 1 | 2.4 | (Skogstad *et al.*, 2006) |
| Hungary | 1 | 2.4 | (Voros *et al.*, 2022) |
| **Publication Years** |  |  |  |
| 1994-2000 | 2 | 4.8 | (Camgan, 1994; Talseth *et al.*, 1999) |
| 2001-2010 | 8 | 19.0 | (Barnes *et al.*, 2002; Cedereke & Öjehagen, 2007; Chung, 2010; Denneson *et al.*, 2010; Pirkis *et al.*, 2001; Raingruber, 2002; Skogstad *et al.*, 2006; Spence *et al.*, 2008) |
| 2011-2020 | 19 | 45.2 | (Adler *et al.*, 2020; Chia-Yi Wu *et al.*, 2012; Cleary, 2017; Hagen *et al.*, 2020; Hausmann-Stabile *et al.*, 2018; Heinsch *et al.*, 2020; Hom *et al.*, 2018; John *et al.*, 2020; Jordan *et al.*, 2012; Jordans *et al.*, 2018; Karras *et al.*, 2017; Kodama *et al.*, 2016; Kuramoto-Crawford *et al.*, 2017; Leavey *et al.*, 2016; McKay & Shand, 2018; Pavulans *et al.*, 2012; Renaud *et al.*, 2014; Thompson *et al.*, 2018; Umubyeyi *et al.*, 2016) |
| 2021 | 7 | 16.7 | (Amin *et al.*, 2021; Ammerman *et al.*, 2021; Ilgen *et al.*, 2021; Jones *et al.*, 2021; Lueck & Poe, 2021; Rheinberger *et al.*, 2021; Saini *et al.*, 2021) |
| 2022 | 3 | 7.1 | (de Lange *et al.*, 2022; Podlogar *et al.*, 2022; Voros *et al.*, 2022) |
| 2023 | 3 | 7.1 | (Barry *et al.*, 2023; Holt *et al.*, 2023; Rainbow *et al.*, 2023) |

### Help-seeking needs

#### Barriers to help-seeking

Included studies most commonly reported individual (n=28) and organizational (n=27) level barriers for help-seeking, followed by interpersonal (n=25), community (n=11) and policy/national level barriers (n=8). See Table 3. for a full list of examples, and the following describes the barriers in order from most common to least common. Many barriers spanned multiple levels of the socio-economic model; however, barriers are examined individually below.

**Table 3.** Barriers and facilitators to help-seeking related to suicide prevention care among individuals who are in contact with mental health services.

| Level | Barriers Count | Citations | Examples | Facilitators Count | Citations | Examples |
| --- | --- | --- | --- | --- | --- | --- |
| Individual | 28 | (Adler *et al.*, 2020; Barry *et al.*, 2023; Camgan, 1994; Chia-Yi Wu *et al.*, 2012; Chung, 2010; Cleary, 2017; Denneson *et al.*, 2010; Hagen *et al.*, 2020; Hausmann-Stabile *et al.*, 2018; Heinsch *et al.*, 2020; Holt *et al.*, 2023; Hom *et al.*, 2018; Ilgen *et al.*, 2021; Jones *et al.*, 2021; Jordan *et al.*, 2012; Karras *et al.*, 2017; Kodama *et al.*, 2016; Kuramoto-Crawford *et al.*, 2017; Leavey *et al.*, 2016; Lueck & Poe, 2021; McKay & Shand, 2018; Pavulans *et al.*, 2012; Pirkis *et al.*, 2001; Podlogar *et al.*, 2022; Saini *et al.*, 2021; Skogstad *et al.*, 2006; Spence *et al.*, 2008; Umubyeyi *et al.*, 2016) | Lack of knowledge, negative attitudes towards help-seeking, negative prior experiences in hospitals leading to distrust in hospitalizations, financial concerns | 16 | (Chia-Yi Wu *et al.*, 2012; Chung, 2010; de Lange *et al.*, 2022; Hagen *et al.*, 2020; Hausmann-Stabile *et al.*, 2018; Hom *et al.*, 2018; Ilgen *et al.*, 2021; Jordan *et al.*, 2012; Karras *et al.*, 2017; Kodama *et al.*, 2016; Leavey *et al.*, 2016; McKay & Shand, 2018; Pavulans *et al.*, 2012; Rainbow *et al.*, 2023; Skogstad *et al.*, 2006; Voros *et al.*, 2022) | Knowledge (knowing where to search for help), positive attitudes toward help-seeking, financial means, self-awareness, coping strategies |
| Interpersonal | 25 | (Adler *et al.*, 2020; Ammerman *et al.*, 2021; Camgan, 1994; Chia-Yi Wu *et al.*, 2012; Chung, 2010; Cleary, 2017; Denneson *et al.*, 2010; Hagen *et al.*, 2020; Hausmann-Stabile *et al.*, 2018; | Negative interaction with healthcare professionals, lack of social connection, lack of understanding of | 22 | (Adler *et al.*, 2020; Ammerman *et al.*, 2021; Barnes *et al.*, 2002; Chia-Yi Wu *et al.*, 2012; Chung, 2010; de Lange *et al.*, 2022; Denneson *et al.*, 2010; | Positive relationship with family and friends, friends and family providing emotional support and encouragement, peer |
|  |  | Heinsch *et al.*, 2020; Holt *et al.*, 2023; Hom *et al.*, 2018; Jones *et al.*, 2021; Jordan *et al.*, 2012; Jordans *et al.*, 2018; Kuramoto-Crawford *et al.*, 2017; Lueck & Poe, 2021; McKay & Shand, 2018; Podlogar *et al.*, 2022; Renaud *et al.*, 2014; Rheinberger *et al.*, 2021; Skogstad *et al.*, 2006; Spence *et al.*, 2008; Talseth *et al.*, 1999), | LGBTQ+ issues by healthcare professionals, negative reactions from informal caregivers impacting self-harm tendencies, fear of burdening friends and family with emotional distress, fear of being perceived differently by peers |  | Hagen *et al.*, 2020; Hausmann-Stabile *et al.*, 2018; Heinsch *et al.*, 2020; Holt *et al.*, 2023; Jones *et al.*, 2021; Jordan *et al.*, 2012; Kodama *et al.*, 2016; McKay & Shand, 2018; Pavulans *et al.*, 2012; Raingruber, 2002; Rheinberger *et al.*, 2021; Skogstad *et al.*, 2006; Spence *et al.*, 2008; Talseth *et al.*, 1999; Umubyeyi *et al.*, 2016) | support from those with similar experiences reducing stigma, positive interactions with healthcare professionals building trust and rapport, text message interventions providing information and reminders |
| Organizational/<br>Institutional | 27 | (Adler *et al.*, 2020; Amin *et al.*, 2021; Camgan, 1994; Cedereke & Öjehagen, 2007; Chia-Yi Wu *et al.*, 2012; Chung, 2010; de Lange *et al.*, 2022; Denneson *et al.*, 2010; Hagen *et al.*, 2020; Hausmann-Stabile *et al.*, 2018; Heinsch *et al.*, 2020; Holt *et al.*, 2023; Hom *et al.*, 2018; John *et al.*, 2020; Jones *et al.*, 2021; Jordan *et al.*, 2012; Lueck & Poe, 2021; McKay & Shand, 2018; Pavulans *et al.*, 2012; Pirkis *et al.*, 2001; Podlogar *et al.*, 2022; Renaud *et al.*, 2014; Rheinberger *et al.*, 2021; Skogstad *et al.*, 2006; Spence *et al.*, 2008; Talseth *et al.*, 1999; Umubyeyi *et al.*, 2016) | Healthcare distrust, dehumanization and feelings of isolation during hospitalization, unmet needs and treatment misalignment with patient expectations, limited control over treatment decisions discouraging seeking help, language barriers, long wait-times | 15 | (Adler *et al.*, 2020; Ammerman *et al.*, 2021; Chung, 2010; Cleary, 2017; de Lange *et al.*, 2022; Denneson *et al.*, 2010; Hagen *et al.*, 2020; Holt *et al.*, 2023; Jordan *et al.*, 2012; Raingruber, 2002; Rheinberger *et al.*, 2021; Skogstad *et al.*, 2006; Spence *et al.*, 2008; Talseth *et al.*, 1999; Umubyeyi *et al.*, 2016) | Holistic care that creates a supportive atmosphere was reported to promote help-seeking, normalization of presence of suicidal ideation and taking time to find out where these thoughts came from, asking relevant and inclusive questions about sexual orientation, gender identity, and related issues was perceived as critical, individualized care, tailored to each person's unique needs and circumstances, follow-up services, including ongoing support and follow-up appointments |
| Community | 11 | (Adler *et al.*, 2020; Amin *et al.*, 2021; Barry *et al.*, 2023; Chung, 2010; Holt *et al.*, 2023; John *et al.*, 2020; Jordan *et al.*, 2012; Kuramoto-Crawford *et al.*, 2017; Leavey *et al.*, 2016; Pavulans *et al.*, 2012; Thompson *et al.*, 2018) | Rurality of residence, lack of accessible services, lack of integration of public health suicide prevention initiatives in primary care settings | 10 | (Adler *et al.*, 2020; Barnes *et al.*, 2002; Chia-Yi Wu *et al.*, 2012; Chung, 2010; Heinsch *et al.*, 2020; Jordan *et al.*, 2012; Leavey *et al.*, 2016; Pavulans *et al.*, 2012; Thompson *et al.*, 2018; Umubyeyi *et al.*, 2016) | Availability of community resources, nearby hospitals, open discussions about mental health and suicide reducing stigma, accessible mental health services, community-based support systems, informal support networks beyond families and friends providing encouragement, crisis hotlines and dissemination of information about assistance resources |
| Policy/National/Law | 8 | (Amin *et al.*, 2021; Chung, 2010; Denneson *et al.*, 2010; Heinsch *et al.*, 2020; Holt *et al.*, 2023; Jordans *et al.*, 2018; Renaud *et al.*, 2014; Umubyeyi *et al.*, 2016). | Lack of health insurance | 1 | (Chung, 2010) | Financial benefits provided through government assistance programs |

##### Individual Barriers

Barriers at the individual level included a lack of knowledge regarding where to seek help, or lack of awareness regarding own mental health (Chung, 2010; Cleary, 2017; Hom *et al*., 2018; Jordan *et al*., 2012; Kodama *et al*., 2016; Kuramoto-Crawford *et al*., 2017) alongside attitudes, often related to prior experiences of support. This included perceptions that support would not be effective (Adler *et al*., 2020; Chia-Yi Wu *et al*., 2012; Cleary, 2017; Heinsch *et al*., 2020; Pavulans *et al*., 2012), and a distrust of mental health services and professionals (Adler *et al*., 2020; Hagen *et al*., 2020; Hausmann-Stabile *et al*., 2018; Holt *et al*., 2023; Jones *et al*., 2021; Skogstad *et al*., 2006; Spence *et al*., 2008). Further individual barriers included fear and apprehension, including worries about hospital presentation and involuntary hospitalization (Adler *et al*., 2020; Holt *et al*., 2023; Jones *et al*., 2021; Jordan *et al*., 2012; Pavulans *et al*., 2012; Rheinberger *et al*., 2021). A study examining the impact of previous mental health care on current disclosure of suicide risk found that men aged over 50 with a history of mental health inpatient care were more likely to explicitly not disclose current suicidal thoughts and behaviours than individuals who received outpatient or no previous care (Podlogar *et al*., 2022), possibly reflecting fear of re-hospitalization. Additional fears include police involvement (Holt *et al*., 2023), and potential loss of career (Adler *et al*., 2020). Apprehension surrounding imposing emotional burden and distress on families was also identified (Chung, 2010; Heinsch *et al*., 2020) where in some cases individuals did not want to seek help in case this resulted in family members discovering their experience with suicidality or other personal information about themselves such as sexuality (Holt *et al*., 2023). Additional individual barriers identified included financial concerns regarding seeking help (e.g., missing work, not being able to afford out-of-pocket costs) (Chung, 2010; Heinsch *et al*., 2020; Holt *et al*., 2023; Hom *et al*., 2018; Kuramoto-Crawford *et al*., 2017), spiritual and cultural beliefs (Chung, 2010), and exacerbating mental health conditions, such as behavioural avoidance (Cleary, 2017; Lueck & Poe, 2021).

##### Organizational Barriers

At the organizational level, prior negative experiences were cited as an important barrier preventing help-seeking. Such experiences included experiences of ‘dehumanization’, ‘alienation’ and ‘isolation’ during hospitalization, and having basic and cultural needs unmet (Adler *et al*., 2020; de Lange *et al*., 2022; Hausmann-Stabile *et al*., 2018; Holt *et al*., 2023; Jones *et al*., 2021; Pirkis *et al*., 2001; Spence *et al*., 2008; Talseth *et al*., 1999, 1999; Umubyeyi *et al*., 2016). Furthermore, multiple studies identified perceived lack of knowledge and understanding among healthcare providers. This included needs related to mental health and suicide prevention (Camgan, 1994; de Lange *et al*., 2022; Rheinberger *et al*., 2021), alongside needs related to individual’s identities, for example sexual and gender identity (de Lange *et al*., 2022). Some samples described being turned away from services, where some patients felt their needs were viewed by services as either ‘not serious enough’ or ‘too complex’ by healthcare providers (Adler *et al*., 2020; McKay & Shand, 2018). Additionally, some samples identified that within emergency departments, physical rather than mental health issues were prioritised (Rheinberger *et al*., 2021; Spence *et al*., 2008). Further organizational barriers identified included language barriers (Chung, 2010), referral mechanisms (Jordan *et al*., 2012), understaffing and long waiting times (Adler *et al*., 2020; Rheinberger *et al*., 2021; Spence *et al*., 2008), the chaotic and rushed nature of settings such as emergency departments and inpatient units (Adler *et al*., 2020; Rheinberger *et al*., 2021; Talseth *et al*., 1999), lack of privacy (Rheinberger *et al*., 2021) and poor assistance post-discharge (Camgan, 1994; Heinsch *et al*., 2020; Rheinberger *et al*., 2021).

##### Interpersonal Barriers

Several interpersonal level barriers were identified as influencing individuals’ willingness to seek help. Stigma was commonly cited as a key barrier to help-seeking. Participants expressed concerns about being perceived as ‘weak’ or ‘weird’ by others (Adler *et al*., 2020; Cleary, 2017; Hausmann-Stabile *et al*., 2018; Heinsch *et al*., 2020; Holt *et al*., 2023; Hom *et al*., 2018; Jordan *et al*., 2012; Umubyeyi *et al*., 2016). This was particularly pronounced among men and some occupational groups such as the military and firefighters.

Furthermore, individuals feared being perceived or treated differently or blamed by peers and colleagues following disclosure of distress (Adler *et al*., 2020; Chia-Yi Wu *et al*., 2012; Hom *et al*., 2018; Jordan *et al*., 2012). Negative interpersonal experiences with medical professionals, marked by a superficial nature, lack of compassion and empathy, alongside sometimes stigmatising views also led to feelings of frustration and in some cases, reluctance to seek professional help (Camgan, 1994; Chia-Yi Wu *et al*., 2012; Hausmann-Stabile *et al*., 2018; Holt *et al*., 2023; Rheinberger *et al*., 2021; Spence *et al*., 2008). Furthermore, while informal caregivers were described as having the potential to provide support, negative reactions from the caregivers were also at times described as contributory to reduced help-seeking behaviours (Spence *et al*., 2008). After a suicide attempt, individuals varied in their willingness to confide in family and friends, reflecting an ongoing hesitation to disclose stressors due to concerns of creating emotional burden (Chung, 2010; Spence *et al*., 2008). However, one study which examined suicide attempt disclosure among veterans found that although many had experienced unhelpful reactions (more commonly from romantic partners and friends), such reactions were not associated with reported likelihood of seeking help for suicidal thoughts or behaviours in the future (Ammerman *et al*., 2021). Finally, some studies identified a lack of social connection or support system as a barrier to support (Hagen *et al*., 2020; Holt *et al*., 2023; Renaud *et al*., 2014; Talseth *et al*., 1999).

##### Community and National/Policy Barriers

Community-level barriers included a lack of resources, such as the absence of nearby hospitals for accessing care (Adler *et al*., 2020; Barry *et al*., 2023; Heinsch *et al*., 2020; Holt *et al*., 2023; Spence *et al*., 2008), and lack of integration of public health suicide prevention initiatives in primary care settings (John *et al*., 2020). For example, individuals with a rural place of residence experienced barriers (such as travel time) that negatively influenced help-seeking behaviours (Barry *et al*., 2023). A study found that refugees in Sweden had lower rates of specialised healthcare both prior to and after a suicide attempt compared to Swedish-born individuals (Amin *et al*., 2021). National/policy-level barriers beyond the community level included lack of health insurance (Chung, 2010; Kuramoto-Crawford *et al*., 2017; Umubyeyi *et al*., 2016), as healthcare coverage is governed at the national or federal level and varies across countries.

#### Facilitators to help-seeking

Facilitators for help-seeking were often identified as the opposite to many of the barriers described above and were also not mutually exclusive. The most commonly reported facilitators in the included studies were within the interpersonal (n=22), individual (n=16), and organizational (n=15) domains, followed by community (n=10) and policy levels (n=1). See Table 3. for a full list of examples, and the following describes the facilitators in order from most common to least common.

##### Interpersonal Facilitators

At the interpersonal level, friends and/or family played a crucial role in facilitating help-seeking by offering emotional support and encouragement to seek help (Adler *et al*., 2020; Chia-Yi Wu *et al*., 2012; Hagen *et al*., 2020; Hausmann-Stabile *et al*., 2018; Holt *et al*., 2023; McKay & Shand, 2018). Sometimes this social support was critical when individuals chose not to, or did not feel able to seek help themselves (Barnes *et al*., 2002; Heinsch *et al*., 2020; Rheinberger *et al*., 2021). In the case of professionals in the army, some stated support from command was instrumental to accessing support, however, some also recognised feeling that they were then treated differently following disclosure (Adler *et al*., 2020). Peer support from those who had shared similar experiences also encouraged help-seeking behaviour and reduced perceived stigma (de Lange *et al*., 2022; Jordans *et al*., 2018). Past positive interactions with healthcare professionals, where individuals had rapport in a trusting, caring, confidential, and respectful relationship motivated service users to seek help or disclose their distress (Adler *et al*., 2020; Chia-Yi Wu *et al*., 2012; Chung, 2010; Hagen *et al*., 2020; Hausmann-Stabile *et al*., 2018; Holt *et al*., 2023; Jones *et al*., 2021; McKay & Shand, 2018; Rheinberger *et al*., 2021; Talseth *et al*., 1999). In one study, veterans rated mental health care professionals as the most helpful person to disclose to regarding their experience of a prior suicide attempt. Among this sample, reported positive reactions to disclosure was positively associated with reported likelihood of disclosing their experience to others in the future, but not with reported likelihood of future help-seeking (Ammerman *et al*., 2021). Positive experiences such as being acknowledged and treated as valued individuals by healthcare professionals, and staff members acknowledging the socio-cultural needs of patients were also facilitators (Chung, 2010; Hagen *et al*., 2020; Holt *et al*., 2023). However one study, which included individuals currently imprisoned, reported that individuals who had no prior contact with a prison psychologist tended to have higher intentions to seek future help for mental health concerns including suicidal thoughts than those with contact (Skogstad *et al*., 2006). Another study described a text messaging intervention that promoted help-seeking behaviours whereby texts including information about social welfare services and reminders about medical appointments were sent (Kodama *et al*., 2016).

##### Individual Facilitators

At the individual level, having positive attitudes or confidence (Ilgen *et al*., 2021; Skogstad *et al*., 2006), capability to search for help from an online source or the community, knowing where to seek help (Ilgen *et al*., 2021; McKay & Shand, 2018; Voros *et al*., 2022), self-awareness of their own mental health symptoms (Chia-Yi Wu *et al*., 2012; Chung, 2010) and desire and willingness to seek help (Rainbow *et al*., 2023; Skogstad *et al*., 2006) acted as facilitators. It is interesting to note that in one study, attitude was the strongest predictor for help-seeking while suicide ideation measured on Suicidal Ideation Questionnaire was not associated with intentions to seek help (Skogstad *et al*., 2006). It was discussed in one study how previous positive experience in mental health services could lead to positive attitudes for future help-seeking (Holt *et al*., 2023). Individual’s sense of autonomy, believing that they are in control of illness, was also important for help-seeking behaviours (Hagen *et al*., 2020). Coping strategies used during crisis or mental health challenges were important in supporting individuals during future help-seeking behaviours (Jordan *et al*., 2012; Rainbow *et al*., 2023).

##### Organizational Facilitators

At the organizational level, holistic and supportive care alongside provision of detailed mental health assessments was reported as promoting help-seeking within mental health services (Hagen *et al*., 2020; Rheinberger *et al*., 2021). In addition, normalization of the presence of suicidal ideation and taking time to find out where these thoughts came from was helpful (de Lange *et al*., 2022). Young adults who identified themselves as sexual and gender minority group members reported that asking relevant and inclusive questions about sexual orientation, gender identity, and related characteristics was perceived as essential when accessing informal or formal mental health care (de Lange *et al*., 2022). Individualized care, tailored to each person’s unique needs and circumstances, also enhanced the likelihood of engagement with mental health services by addressing specific challenges. Follow-up services, including ongoing support and follow-up appointments, reinforced the idea that individuals are valued and cared for beyond their initial crisis, encouraging continued engagement with mental health resources (Adler *et al*., 2020; Cleary, 2017; Holt *et al*., 2023; Jordan *et al*., 2012).

##### Community Facilitators

At the community level, a social environment where mental health and suicide are openly discussed can reduce stigma and normalize help-seeking behaviours (Chia-Yi Wu *et al*., 2012; Raingruber, 2002). Having easily accessible mental health services encourage individuals to seek help (Umubyeyi *et al*., 2016). Access to community-based support systems (Jordan *et al*., 2012), including community centers, mental health education programs, and support groups foster a sense of belonging and social interaction, which breaks down barriers to seeking help. Informal support networks consisting of neighbours, and even ‘strangers’ provide emotional support and encouragement for individuals to seek help (Chung, 2010). Moreover, the promotion of crisis hotlines (Karras *et al*., 2017), along with disseminating information about available assistance resources was one of the ways to increase individuals’ knowledge of where to go when in need of help. While rurality of residence was reported as a barrier to help seeking behaviours due to limited access to services, rurality was reported as one predictor of using hotline services (Thompson *et al*., 2018). At the national level, a study identified the accessibility of financial benefits as a facilitative factor. These benefits played a crucial role in alleviating participants’ stressors and enabling them to take time off from work, as directed by their mental health providers (Chung, 2010).

## Discussion

This scoping review identified 42 empirical studies that described the suicide-related help-seeking needs of individuals who are in contact with and/or accessing mental health services in a range of healthcare settings, such as in-patient, outpatient, emergency, and crisis line services. The majority of articles were published between 2011 and 2020, and 13 articles were published within the past three years (2021-2023). Almost all studies focused on suicide attempts and/or ideation as suicide phenomena. Authors commonly described characteristics of participants’ age, sex, marital status, and gender. Barriers and facilitators to help-seeking behaviors related to suicide prevention care among individuals in contact with mental health services were frequently reported at individual, interpersonal and organizational levels. These barriers encompass a range of factors, including stigma, lack of awareness, and systemic deficiencies in healthcare provision. Conversely, facilitators include supportive relationships, positive attitudes, accessible services, community resources, and financial benefits.

An important observation from this review when examining participant-reported characteristics is the presence of reported indigeneity in the included studies. The current review focused on individuals who are in contact with health services to receive support for suicide-related thoughts and behaviors, and it is noteworthy that we found limited studies specific to Indigenous peoples among other racialized groups. There was one study from New Zealand that reported indigeneity, aiming to examine the help-seeking behaviors of prisoners seeking assistance from a prison psychologist for their suicidal feelings (Skogstad *et al*., 2006). Indigenous peoples are at a higher risk of suicide, and these heightened risks are attributed to systemic marginalization, and the enduring effects of cultural and social disparities, all of which are deeply rooted in historical colonization (Pollock *et al*., 2018). In addition, their experiences with health services impact their help-seeking behaviors, and some may avoid hospitals due to lack of trust (Coombes *et al*., 2022; Mbuzi *et al*., 2017). This observation may explain the scarcity of studies examining the help-seeking behaviors of Indigenous peoples who are already accessing care, positing important areas for further investigation in future research.

While the included studies lacked extensive discussion on how barriers and varying intersectional identities affect individuals differently, it is crucial to consider the intersectional perspective when interpreting the findings of this review. It is important to be mindful that some individuals may encounter more barriers when seeking help compared to others. Individuals who self-identified as sexual and gender minorities, relationships with peers and/or families, and asking inclusive questions played a critical role in facilitating help-seeking behaviours (de Lange *et al*., 2022). This finding highlights the need to ensure practitioners’ competency for working with diverse groups. Middle-aged men, especially those working in ’masculine’ occupations such as the military, reported fear of being perceived as ‘weak’ as a barrier for seeking help (Adler *et al*., 2020; Cleary, 2017). In addition, disclosing mental health needs can be more challenging in certain cultures due to the presence of a more pronounced stigma (Chung, 2010). Social attitudes towards mental health and suicide have made significant progress over the past decades, yet it is important to acknowledge that lingering stigma surrounding suicide and mental health still persists. The normalization of help-seeking is critical not only for men but for everyone. Healthcare institutions and the broader social environment should be encouraged to work collaboratively to provide safe and non-stigmatized spaces targeting marginalized groups with the aim of supporting and normalizing help seeking. Recommendations for anti-stigma include training healthcare professionals (Henderson *et al*., 2014; Sreeram *et al*., 2022) and launching public awareness campaigns (Pirkis *et al*., 2019) to reach a wider societal audience.

The current literature also describes barriers to help-seeking from the perspectives of parents and healthcare professionals, and they provide valuable evidence relevant to the understanding of the findings of this review (Moskos *et al*., 2007; O’Keeffe *et al*., 2021). Parents’ perceived barriers to help-seeking for their children included hopelessness, fear of being seen as ‘weak’, and feeling apprehensive about disclosing mental health needs (Moskos *et al*., 2007). In addition, carers reported perceived feelings of judgment experienced by service users when seeking mental health help in an emergency department (Moskos *et al*., 2007). Seeking mental health help in highly medicalized settings, such as an emergency department, made both caregivers and service users feel as if they were being judged and perceived as ‘time wasters’, using resources unnecessarily, or being considered less worthy than those with physical health issues (Moskos *et al*., 2007). A need for human connection and therapeutic interactions in healthcare setting was reported (Moskos *et al*., 2007), which is consistent with our review findings of facilitators for help-seeking for individuals experiencing suicidal ideation and behaviours. Emergency department healthcare professionals also recognized the therapeutic value of a person feeling listened to; however, it was reported that the emergency department was not the right environment for therapeutic conversations (O’Keeffe *et al*., 2021). This is also described elsewhere in the literature and explained that traditional emergency practice is deeply rooted in biomedical practice which may conflict with expectations of patients seeking mental health support (Shin *et al*., 2020). For example, patients described formulaic question-and-answer risk assessments, which is often the way emergency department professionals practice, as an unhelpful way of addressing their needs; however, healthcare professionals reported feeling safer using such tools as they are fearful of blame if someone goes on to die by suicide (O’Keeffe *et al*., 2021). It is important to acknowledge that emergency department professionals have the intention to assist, yet they often feel ‘powerless’ (O’Keeffe *et al*., 2021). Emergency department professionals also recognized the complex nature of suicide ideation and behavior, yet they did not hold the belief that they could adequately address someone’s needs or effectively help them cease self-harm (O’Keeffe *et al*., 2021).

A few studies were excluded from this review due to an ineligible context, as individuals were not in contact with health services seeking or accessing mental health care. These studies nonetheless provide valuable evidence relevant to the findings of this review. It was found that individuals presented to the emergency department less frequently during the pandemic for suicide ideation (Sass *et al*., 2022; Sveticic *et al*., 2021). Participants described feeling like a burden to others due to an awareness of all healthcare services struggling under the pressure of COVID-19 (Sass *et al*., 2022). Digital tools can serve as a valuable tool to address the help-seeking needs of individuals, particularly those who are hesitant to visit hospitals, especially amidst pandemic situations and where there are concerns about being judged. Technologies offer a variety of platforms and solutions that enable individuals to seek help remotely, thereby minimizing the barriers linked to physical presence. Virtual mental health platforms, telemedicine services, and crisis helplines accessible via phone or text provide discreet channels for seeking assistance (Rassy *et al*., 2021; Shin *et al*., 2023). As demonstrated by the current review’s findings, communication through crisis lines was found to be helpful in disclosing mental health concerns and addressing needs (Ilgen *et al*., 2021). These types of technologies not only preserve anonymity but also allow individuals to communicate from the comfort of their own environment, thus alleviating the fear of judgment often associated with face-to-face interactions (Lehtimaki *et al*., 2021).

The frequency counts are not appropriate to infer hierarchy of barriers and facilitators we identified in this review. Additionally, this review did not perform meta-analysis to determine the overall effect of these barriers and facilitators of help-seeking behaviours. However, it is worth noting that barriers and facilitators span all levels, highlighting important considerations for informing suicide prevention efforts at multiple levels to remove barriers and promote facilitators. Technologies and digital tools can augment suicide prevention care in clinical settings and online assessments are often preferred and reliable means for assessing sensitive information such as suicidal thoughts and behaviors (Iorfino *et al*., 2017). As we plan our next phase of the overarching project of informing the next iteration of a digital tool to support suicide prevention care, it is critical to consider the identified barriers to help-seeking and the existing discrepancies in the adoption of digital tools. As such, one recommendation to address complex barriers such as stigma without exacerbating existing digital equity gaps is to work closely with individuals with lived experience to identify appropriate and effective interventions (Lyles *et al*., 2023). Also, when looking at the existing apps for suicide prevention, 66 apps were identified from app stores, of which 35 apps were found in both iOS and Android app stores (Wilks *et al*., 2021). These apps were assessed for their contents, such as the presence of a safety plan and crisis line access (Wilks *et al*., 2021). However, it is also important to consider the list of barriers and facilitators outlined in Table 3 as design considerations where appropriate, including the use of inclusive language, fostering trust and transparency regarding data use, and ensuring the app is freely accessible. This approach is essential to prevent the replication of barriers to help-seeking using digital solutions.

Despite the valuable insights obtained from this rapid scoping review, limitations should be acknowledged. The expedited nature of the review process inherently led to trade-offs that might impact the comprehensiveness of findings. Without a quality assessment of included studies, we likely introduced studies of varied methodological quality. Additionally, the accelerated process led to the omission of grey literature searches, and search results may not have been as exhaustive as in other types of literature reviews (e.g., systematic). Finally, the review included solely English-language articles. This might have introduced publication bias and may not be representative of help-seeking behaviors and needs in non-English speaking countries.

## Conclusion

This review outlines the characteristics of studies that describe the suicide-related help-seeking needs of individuals in contact with mental health services in any healthcare settings. The majority of included studies were observational or qualitative in nature, varying in their details about the barriers and facilitators for seeking mental health support for suicide ideation and behaviors. Barriers and facilitators were detected across all levels of the adapted socio-ecological model. Reported barriers and facilitators were not isolated to one single level; instead, they interacted, underscoring the intricate and interrelated nature of help-seeking. Suggestions to address help-seeking needs identified by studies include use of inclusive language, fostering trust, and normalizing help-seeking behaviours. However, it is important to recognise that certain barriers, such as stigma, are deeply rooted and systemic in nature, representing challenges that have persisted for many years. As our research team moves forward with designing a new iteration of a suicide prevention digital tool, it is crucial to involve individuals with lived experience. This engagement will help identify and address barriers where appropriate and capitalize on facilitators.

## Data Availability

All data produced in the present study are available upon reasonable request to the authors

## Acknowledgements

We would like to thank librarian, Reena Besa, who helped us build the search strategy.

## References

Adams, C., Gringart, E., & Strobel, N. (2022) Explaining adults’ mental health help-seeking through the lens of the theory of planned behavior: a scoping review. Systematic Reviews, 11, 160. doi:10.1186/s13643-022-02034-y

Adler, A., Jager-Hyman, S., Brown, G. K., et al. (2020) A qualitative investigation of barriers to seeking treatment for suicidal thoughts and behaviors among army soldiers with a deployment history. Archives of Suicide Research, 24, 251–268. doi:10.1080/13811118.2019.1624666

Aguirre Velasco, A., Cruz, I. S. S., Billings, J., et al. (2020) What are the barriers, facilitators and interventions targeting help-seeking behaviours for common mental health problems in adolescents? A systematic review. BMC Psychiatry, 20, 293. doi:10.1186/s12888-020-02659-0

Amin, R., Rahman, S., Tinghog, P., et al. (2021) Healthcare use before and after suicide attempt in refugees and Swedish-born individuals. Social Psychiatry and Psychiatric Epidemiology: The International Journal for Research in Social and Genetic Epidemiology and Mental Health Services, 56, 325–338. doi:10.1007/s00127-020-01902-z

Ammerman, B. A., Carter, S. P., Gebhardt, H. M., et al. (2021) An initial investigation of suicide attempt disclosures among US veterans. Crisis: The Journal of Crisis Intervention and Suicide Prevention, 42, 411–417. doi:10.1027/0227-5910/a000727

Barnes, Ikeda, & Kresnow (2002) Help-seeking behavior prior to nearly lethal suicide attempts. Suicide and Life-Threatening Behavior, 32, 68–75. doi:10.1521/suli.32.1.5.68.24217

Barry, R., Rehm, J., de Oliveira, C., et al. (2023) Help-seeking behavior among adults who attempted or died by suicide in Ontario, Canada. Suicide & Life-Threatening Behavior, 53, 54–63. doi:10.1111/sltb.12921

Bergqvist, E., Probert-Lindström, S., Fröding, E., et al. (2022) Health care utilisation two years prior to suicide in Sweden: a retrospective explorative study based on medical records. BMC Health Services Research, 22, 664.

Biswas, T., Scott, J. G., Munir, K., et al. (2020) Global variation in the prevalence of suicidal ideation, anxiety and their correlates among adolescents: A population based study of 82 countries. eClinicalMedicine, 24. doi:10.1016/j.eclinm.2020.100395

Borges, G., Nock, M. K., Abad, J. M. H., et al. (2010) Twelve-month prevalence of and risk factors for suicide attempts in the World Health Organization World Mental Health Surveys. The Journal of Clinical Psychiatry, 71, 21777.

Cairns, J.-M., Graham, E., & Bambra, C. (2017) Area-level socioeconomic disadvantage and suicidal behaviour in Europe: a systematic review. Social Science & Medicine, 192, 102– 111.

Camgan, J. T. (1994) The psychosocial needs of patients who have attempted suicide by overdose. Journal of Advanced Nursing, 20, 635–642. doi:10.1046/j.1365-2648.1994.20040635.x

Cassidy, S. A., Robertson, A., Townsend, E., et al. (2020) Advancing our understanding of self-harm, suicidal thoughts and behaviours in autism. Journal of Autism and Developmental Disorders, 50, 3445–3449.

Cedereke, M., & Öjehagen, A. (2007) Formal and informal help during the year after a suicide attempt: A one-year follow-up. International Journal of Social Psychiatry, 53, 419–429. doi:10.1177/0020764007078345

Chia-Yi Wu, Whitley, R., Stewart, R., et al. (2012) Pathways to Care and Help-Seeking Experience Prior to Self-Harm: A Qualitative Study in Taiwan. Journal of Nursing Research (Lippincott Williams & Wilkins), 20, 32–42. doi:10.1097/JNR.0b013e3182466e64

Chung, I. (2010) Changes in the sociocultural reality of chinese immigrants: challenges and opportunities in help-seeking behaviour. The International Journal of Social Psychiatry, 56, 436–47. doi:10.1177/0020764009105647

Cleary, A. (2017) Help-seeking patterns and attitudes to treatment amongst men who attempted suicide. Journal of Mental Health (Abingdon, England), 26, 220–224. doi:10.3109/09638237.2016.1149800

Clements, C., Hawton, K., Geulayov, G., et al. (2019) Self-harm in midlife: analysis using data from the Multicentre Study of Self-harm in England. The British Journal of Psychiatry, 215, 600–607.

Coombes, J., Hunter, K., Bennett-Brook, K., et al. (2022) Leave events among Aboriginal and Torres Strait Islander people: a systematic review. BMC Public Health, 22, 1488. doi:10.1186/s12889-022-13896-1

Cornally, N., & McCarthy, G. (2011) Help seeking behaviour: A concept analysis. International Journal of Nursing Practice, 17, 280–288.

Covidence systematic review software (2019). Melbourne, Australia. Retrieved from www.covidence.org

Crenshaw, K. (1989) Demarginalizing the intersection of race and sex: A black feminist critique of antidiscrimination doctrine, feminist theory and antiracist politics. U. Chi. Legal f., 139.

Crenshaw, K. (1991) Mapping the Margins: Intersectionality, Identity Politics, and Violence against Women of Color. Stanford Law Review, 43, 1241–1299. doi:10.2307/1229039

de Lange, J., van Bergen, D. D., Baams, L., et al. (2021) Experiences and needs of sexual and gender minority young adults with a history of suicidal ideation regarding formal and informal mental healthcare. Sexuality Research & Social Policy: A Journal of the NSRC, No-Specified. doi:10.1007/s13178-021-00657-9

de Lange, J., van Bergen, D. D., Baams, L., et al. (2022) Experiences and Needs of Sexual and Gender Minority Young Adults with a History of Suicidal Ideation Regarding Formal and Informal Mental Healthcare. Sexuality Research and Social Policy, 19, 1829–1841. doi:10.1007/s13178-021-00657-9

De Leo, D., Goodfellow, B., Silverman, M., et al. (2021) International study of definitions of English-language terms for suicidal behaviours: a survey exploring preferred terminology. BMJ Open, 11, e043409.

Denneson, Basham C., Dickinson K.C., et al. (2010) Suicide risk assessment and content of VA health care contacts before suicide completion by veterans in Oregon. Psychiatric Services, 61, 1192–1197. doi:10.1176/ps.2010.61.12.1192

Favril, L., Yu, R., Geddes, J. R., et al. (2023) Individual-level risk factors for suicide mortality in the general population: an umbrella review. The Lancet Public Health, 8, e868–e877.

Favril, L., Yu, R., Uyar, A., et al. (2022) Risk factors for suicide in adults: systematic review and meta-analysis of psychological autopsy studies. BMJ Ment Health, 25, 148–155.

Garritty, C., Gartlehner, G., Nussbaumer-Streit, B., et al. (2021) Cochrane Rapid Reviews Methods Group offers evidence-informed guidance to conduct rapid reviews. Journal of Clinical Epidemiology, 130, 13–22.

Gulliver, A., Griffiths, K. M., & Christensen, H. (2010) Perceived barriers and facilitators to mental health help-seeking in young people: a systematic review. BMC Psychiatry, 10, 113. doi:10.1186/1471-244X-10-113

Hagen, J., Loa Knizek, B., & Hjelmeland, H. (2020) “ … I felt completely stranded”: liminality and recognition of personhood in the experiences of suicidal women admitted to psychiatric hospital. International Journal of Qualitative Studies on Health and Well-Being, 15. doi:10.1080/17482631.2020.1731995

Hausmann-Stabile, C., Gulbas, L., & Zayas, L. H. (2018) Treatment Narratives of Suicidal Latina Teens. Archives of Suicide Research : Official Journal of the International Academy for Suicide Research, 22, 165–172. doi:10.1080/13811118.2017.1304305

Hawton, K., Bale, L., Brand, F., et al. (2020) Mortality in children and adolescents following presentation to hospital after non-fatal self-harm in the multicentre study of self-harm: a prospective observational cohort study. The Lancet Child & Adolescent Health, 4, 111– 120.

Hawton, K., Bergen, H., Cooper, J., et al. (2015) Suicide following self-harm: findings from the multicentre study of self-harm in England, 2000–2012. Journal of Affective Disorders, 175, 147–151.

Heinsch, M., Sampson, D., Huens, V., et al. (2020) Understanding ambivalence in help-seeking for suicidal people with comorbid depression and alcohol misuse. PLoS ONE, 15. doi:10.1371/journal.pone.0231647

Henderson, C., Noblett, J., Parke, H., et al. (2014) Mental health-related stigma in health care and mental health-care settings. The Lancet Psychiatry, 1, 467–482. doi:10.1016/S2215-0366(14)00023-6

Holt, N. R., Botelho, E., Wolford-Clevenger, C., et al. (2023) Previous mental health care and help-seeking experiences: Perspectives from sexual and gender minority survivors of near-fatal suicide attempts. Psychological Services, No-Specified. doi:10.1037/ser0000745

Hom, M. A., Stanley, I. H., Spencer-Thomas, S., et al. (2018) Mental health service use and help-seeking among women firefighters with a career history of suicidality. Psychological Services, 15, 316–324. doi:10.1037/ser0000202

Hsieh, H.-F., & Shannon, S. E. (2005) Three Approaches to Qualitative Content Analysis. Qualitative Health Research, 15, 1277–1288. doi:10.1177/1049732305276687

Ilgen, M. A., Stewart, H. J., Lhermitte, S. L., et al. (2021) Developing and testing crisis line facilitation to encourage help seeking in adults receiving inpatient treatment for a suicidal crisis. Cognitive and Behavioral Practice, 28, 15–21. doi:10.1016/j.cbpra.2020.05.004

Iorfino, F., Davenport, T. A., Ospina-Pinillos, L., et al. (2017) Using New and Emerging Technologies to Identify and Respond to Suicidality Among Help-Seeking Young People: A Cross-Sectional Study. J Med Internet Res, 19, e247. doi:10.2196/jmir.7897

Jang, Y., Kim, G., Hansen, L., et al. (2007) Attitudes of older Korean Americans toward mental health services. Journal of the American Geriatrics Society, 55, 616–620.

John, A., DelPozo-Banos, M., Gunnell, D., et al. (2020) Contacts with primary and secondary healthcare prior to suicide: Case-control whole-population-based study using person-level linked routine data in Wales, UK, 2000-2017. The British Journal of Psychiatry, 217, 717–724. doi:10.1192/bjp.2020.137

Jones, N., Gius, B. K., Shields, M., et al. (2021) Investigating the impact of involuntary psychiatric hospitalization on youth and young adult trust and help-seeking in pathways to care. Social Psychiatry and Psychiatric Epidemiology: The International Journal for Research in Social and Genetic Epidemiology and Mental Health Services, 56, 2017– 2027. doi:10.1007/s00127-021-02048-2

Jordan, J., McKenna, H., Keeney, S., et al. (2012) Providing meaningful care: Learning from the experiences of suicidal young men. Qualitative Health Research, 22, 1207–1219. doi:10.1177/1049732312450367

Jordans, M., Rathod, S., Fekadu, A., et al. (2018) Suicidal ideation and behaviour among community and health care seeking populations in five low- and middle-income countries: A cross-sectional study. Epidemiology and Psychiatric Sciences, 27, 393–402. doi:10.1017/S2045796017000038

Karras, E., Lu, N., Elder, H., et al. (2017) Promoting help seeking to veterans: A comparison of public messaging strategies to enhance the use of the veterans crisis line. Crisis: The Journal of Crisis Intervention and Suicide Prevention, 38, 53–62. doi:10.1027/0227-5910/a000418

Kidd, G., Marston, L., Nazareth, I., et al. (2023) Suicidal thoughts, suicide attempt and non-suicidal self-harm amongst lesbian, gay and bisexual adults compared with heterosexual adults: analysis of data from two nationally representative English household surveys. Social Psychiatry and Psychiatric Epidemiology. doi:10.1007/s00127-023-02490-4

Knipe, D., Padmanathan, P., Newton-Howes, G., et al. (2022) Suicide and self-harm. The Lancet, 399, 1903–1916.

Kodama, T., Syouji, H., Takaki, S., et al. (2016) Text Messaging for Psychiatric Outpatients: Effect on Help-Seeking and Self-Harming Behaviors. Journal of Psychosocial Nursing and Mental Health Services, 54, 31–7. doi:10.3928/02793695-20160121-01

Kuramoto-Crawford, S. J., Han B., & McKeon R.T. (2017) Self-reported reasons for not receiving mental health treatment in adults with serious suicidal thoughts. Journal of Clinical Psychiatry, 78, e631–e637. doi:10.4088/JCP.16m10989

Leavey, G., Rosato, M., Galway, K., et al. (2016) Patterns and predictors of help-seeking contacts with health services and general practitioner detection of suicidality prior to suicide: a cohort analysis of suicides occurring over a two-year period. BMC Psychiatry, 16, 120. doi:10.1186/s12888-016-0824-7

Lehtimaki, S., Martic, J., Wahl, B., et al. (2021) Evidence on Digital Mental Health Interventions for Adolescents and Young People: Systematic Overview. JMIR Ment Health, 8, e25847. doi:10.2196/25847

Lim, K.-S., Wong, C. H., McIntyre, R. S., et al. (2019) Global lifetime and 12-month prevalence of suicidal behavior, deliberate self-harm and non-suicidal self-injury in children and adolescents between 1989 and 2018: a meta-analysis. International Journal of Environmental Research and Public Health, 16, 4581.

Liu, R. T., Sheehan, A. E., Walsh, R. F. L., et al. (2019) Prevalence and correlates of non-suicidal self-injury among lesbian, gay, bisexual, and transgender individuals: A systematic review and meta-analysis. Clinical Psychology Review, 74, 101783. doi:10.1016/j.cpr.2019.101783

Lovero, K. L., Dos Santos, P. F., Come, A. X., et al. (2023) Suicide in global mental health. Current Psychiatry Reports, 25, 255–262.

Lueck, J. A., & Poe, M. (2021) Bypassing the waitlist: examining barriers and facilitators of help-line utilization among college students with depression symptoms. Journal of Mental Health, 30, 308–314. doi:10.1080/09638237.2020.1760225

Lyles, C. R., Nguyen, O. K., Khoong, E. C., et al. (2023) Multilevel Determinants of Digital Health Equity: A Literature Synthesis to Advance the Field. Annual Review of Public Health, 44, 383–405. doi:10.1146/annurev-publhealth-071521-023913

Mann, J. J., Apter, A., Bertolote, J., et al. (2005) Suicide Prevention StrategiesA Systematic Review. JAMA, 294, 2064–2074. doi:10.1001/jama.294.16.2064

Marconi, E., Monti, L., Marfoli, A., et al. (2023) A systematic review on gender dysphoria in adolescents and young adults: focus on suicidal and self-harming ideation and behaviours. Child and Adolescent Psychiatry and Mental Health, 17, 110. doi:10.1186/s13034-023-00654-3

Martinez, A. B., Co, M., Lau, J., et al. (2020) Filipino help-seeking for mental health problems and associated barriers and facilitators: a systematic review. Social Psychiatry and Psychiatric Epidemiology, 55, 1397–1413. doi:10.1007/s00127-020-01937-2

Mbuzi, V., Fulbrook, P., & Jessup, M. (2017) Indigenous peoples’ experiences and perceptions of hospitalisation for acute care: A metasynthesis of qualitative studies. International Journal of Nursing Studies, 71, 39–49.

McGowan, J., Sampson, M., Salzwedel, D. M., et al. (2016) PRESS Peer Review of Electronic Search Strategies: 2015 Guideline Statement. Journal of Clinical Epidemiology, 75, 40–46. doi:10.1016/j.jclinepi.2016.01.021

McKay, K., & Shand, F. (2018) Advocacy and luck: Australian healthcare experiences following a suicide attempt. Death Studies, 42, 392–399. doi:10.1080/07481187.2017.1359218

McLeroy, K. R., Bibeau, D., Steckler, A., et al. (1988) An Ecological Perspective on Health Promotion Programs. Health Education Quarterly, 15, 351–377.

Miranda-Mendizabal, A., Castellví, P., Parés-Badell, O., et al. (2019) Gender differences in suicidal behavior in adolescents and young adults: systematic review and meta-analysis of longitudinal studies. International Journal of Public Health, 64, 265–283. doi:10.1007/s00038-018-1196-1

Mojtabai, R. (2009) Unmet need for treatment of major depression in the United States. Psychiatric Services, 60, 297–305.

Moskos, M. A., Olson, L., Halbern, S. R., et al. (2007) Utah youth suicide study: Barriers to mental health treatment for adolescents. Suicide and Life-Threatening Behavior, 37, 179– 186.

Murphy, E., Kapur, N., Webb, R., et al. (2012) Risk factors for repetition and suicide following self-harm in older adults: multicentre cohort study. The British Journal of Psychiatry, 200, 399–404.

Nuij, C., van Ballegooijen, W., de Beurs, D., et al. (2021) Safety planning-type interventions for suicide prevention: meta-analysis. British Journal of Psychiatry, 1–8. doi:10.1192/bjp.2021.50

O’Keeffe, S., Suzuki, M., Ryan, M., et al. (2021) Experiences of care for self-harm in the emergency department: Comparison of the perspectives of patients, carers and practitioners. BJPsych Open, 7. doi:10.1192/bjo.2021.1006

Pavulans, K. S., BolmsjÖ, I., Edberg, A.-K., et al. (2012) Being in want of control: Experiences of being on the road to, and making, a suicide attempt. International Journal of Qualitative Studies on Health & Well-Being, 7, 1–11. doi:10.3402/qhw.v7i0.16228

Peters, M. D. J., Godfrey, C. M., Khalil, H., et al. (2015) Guidance for conducting systematic scoping reviews. JBI Evidence Implementation, 13. Retrieved from https://journals.lww.com/ijebh/fulltext/2015/09000/guidance_for_conducting_systematic_scoping_reviews.5.aspx

Peters, M. D. J., Marnie, C., Tricco, A. C., et al. (2020) Updated methodological guidance for the conduct of scoping reviews. JBI Evidence Synthesis, 18. 10.11124/JBIES-20-00167

Pirkis, J., Burgess, P., Meadows, G., et al. (2001) Self-reported needs for care among persons who have suicidal ideation or who have attempted suicide. Psychiatric Services (Washington, D.C.), 52, 381–3.

Pirkis, J., Rossetto, A., Nicholas, A., et al. (2019) Suicide Prevention Media Campaigns: A Systematic Literature Review. Health Communication, 34, 402–414. doi:10.1080/10410236.2017.1405484

Podlogar, M. C., Gutierrez, P. M., & Joiner, T. E. (2022) Past levels of mental health intervention and current nondisclosure of suicide risk among men older than age 50. Assessment, 29, 1611–1621. doi:10.1177/10731911211023577

Pollock, N. J., Naicker, K., Loro, A., et al. (2018) Global incidence of suicide among Indigenous peoples: a systematic review. BMC Medicine, 16, 145. doi:10.1186/s12916-018-1115-6

Qian, J., Zeritis, S., Larsen, M., et al. (2023) The application of spatial analysis to understanding the association between area-level socio-economic factors and suicide: a systematic review. Social Psychiatry and Psychiatric Epidemiology, 58, 843–859. doi:10.1007/s00127-023-02441-z

Qin, P., Syeda, S., Canetto, S. S., et al. (2022) Midlife suicide: A systematic review and meta-analysis of socioeconomic, psychiatric and physical health risk factors. Journal of Psychiatric Research.

Radez, J., Reardon, T., Creswell, C., et al. (2021) Why do children and adolescents (not) seek and access professional help for their mental health problems? A systematic review of quantitative and qualitative studies. European Child & Adolescent Psychiatry, 30, 183–211. doi:10.1007/s00787-019-01469-4

Rainbow, Tatnell R., Blashki G., et al. (2023) Revisiting the factor structure of the suicide-related coping scale: Results from two samples of Australian online help-seekers. Psychiatry Research, 324, 115195. doi:10.1016/j.psychres.2023.115195

Raingruber, B. (2002) Client and provider perspectives regarding the stigma of and nonstigmatizing interventions for depression. Archives of Psychiatric Nursing, 16, 201–7.

Randles, R., & Finnegan, A. (2022) Veteran help-seeking behaviour for mental health issues: a systematic review. BMJ Mil Health, 168, 99–104.

Rassy, J., Bardon, C., Dargis, L., et al. (2021) Information and communication technology use in suicide prevention: Scoping review. Journal of Medical Internet Research, 23, e25288.

Renaud, J., Seguin, M., Lesage, A. D., et al. (2014) Service use and unmet needs in youth suicide: a study of trajectories. Canadian Journal of Psychiatry. Revue Canadienne de Psychiatrie, 59, 523–30.

Rheinberger, Macdonald D., McGillivray L., et al. (2021) “A sustained, productive, constructive relationship with someone who can help”-A qualitative exploration of the experiences of help seekers and support persons using the emergency department during a suicide crisis. International Journal of Environmental Research and Public Health, 18, 10262. doi:10.3390/ijerph181910262

Rudes, G., & Fantuzzi, C. (2022) The association between racism and suicidality among young minority groups: a systematic review. Journal of Transcultural Nursing, 33, 228–238.

Saini, P., Chopra, J., Hanlon, C. A., et al. (2021) A case series study of help-seeking among younger and older men in suicidal crisis. International Journal of Environmental Research and Public Health, 18. doi:10.3390/ijerph18147319

Sass, C., Farley, K., & Brennan, C. (2022) “They have more than enough to do than patch up people like me.” Experiences of seeking support for self harm in lockdown during the COVID 19 pandemic. Journal of Psychiatric & Mental Health Nursing (John Wiley & Sons, Inc.), 29, 544–554. doi:10.1111/jpm.12834

Shin, H. D., Durocher, K., Sequeira, L., et al. (2023) Information and communication technology-based interventions for suicide prevention implemented in clinical settings: a scoping review. BMC Health Services Research, 23, 281. doi:10.1186/s12913-023-09254-5

Shin, H. D., Price, S., & Aston, M. (2020) A poststructural analysis: Current practices for suicide prevention by nurses in the emergency department and areas of improvement. Journal of Clinical Nursing, n/a. doi:10.1111/jocn.15502

Skogstad, Deane F.P., & Spicer J. (2006) Social-cognitive determinants of help-seeking for mental health problems among prison inmates. Criminal Behaviour and Mental Health, 16, 43–59. doi:10.1002/cbm.54

Spence, J. M., Bergmans, Y., Strike, C., et al. (2008) Experiences of substance-using suicidal males who present frequently to the emergency department. CJEM, 10, 339–346. doi:10.1017/s1481803500010344

Sreeram, A., Cross, W. M., & Townsin, L. (2022) Anti-stigma initiatives for mental health professionals—A systematic literature review. Journal of Psychiatric and Mental Health Nursing, 29, 512–528. doi:10.1111/jpm.12840

Stene-Larsen, K., & Reneflot, A. (2019) Contact with primary and mental health care prior to suicide: A systematic review of the literature from 2000 to 2017. Scandinavian Journal of Public Health, 47, 9–17. doi:10.1177/1403494817746274

Sveticic, J., Stapelberg, N. J. C., & Turner, K. (2021) Suicide prevention during COVID-19: identification of groups with reduced presentations to emergency departments. Australasian Psychiatry, 29, 333–336. doi:10.1177/1039856221992632

Talseth, A.-G., Lindseth, A., Jacobsson, L., et al. (1999) The meaning of suicidal psychiatric in-patients’ experiences of being cared for by mental health nurses. Journal of Advanced Nursing, 29, 1034–1041. doi:10.1046/j.1365-2648.1999.00990.x

Thompson, L. K., Sugg, M. M., & Runkle, J. R. (2018) Adolescents in crisis: A geographic exploration of help-seeking behavior using data from Crisis Text Line. Social Science & Medicine (1982), 215, 69–79. doi:10.1016/j.socscimed.2018.08.025

Tricco, A. C., Antony, J., Zarin, W., et al. (2015) A scoping review of rapid review methods. BMC Medicine, 13, 224. doi:10.1186/s12916-015-0465-6

Tricco, A. C., Lillie, E., Zarin, W., et al. (2018) PRISMA Extension for Scoping Reviews (PRISMA-ScR): Checklist and Explanation. Annals of Internal Medicine, 169, 467–473. doi:10.7326/M18-0850

Umubyeyi, A., Mogren, I., Ntaganira, J., et al. (2016) Help-seeking behaviours, barriers to care and self-efficacy for seeking mental health care: a population-based study in Rwanda. Social Psychiatry and Psychiatric Epidemiology, 51, 81–92. doi:10.1007/s00127-015-1130-2

Van Meter, A. R., Knowles, E. A., & Mintz, E. H. (2022) Systematic Review and Meta-analysis: International Prevalence of Suicidal Ideation and Attempt in Youth. Journal of the American Academy of Child & Adolescent Psychiatry. doi:10.1016/j.jaac.2022.07.867

Voros, Fekete S., Szabo Z., et al. (2022) High prevalence of suicide-related internet use among patients with depressive disorders - a cross-sectional study with psychiatric in-patients. Psychiatry Research, 317, 114815. doi:10.1016/j.psychres.2022.114815

WHO (2014) Preventing suicide: A global imperative. World Health Organization. Retrieved June 27, 2021, from https://www.who.int/publications/i/item/9789241564779

Wilks, C. R., Chu, C., Sim, D., et al. (2021) User Engagement and Usability of Suicide Prevention Apps: Systematic Search in App Stores and Content Analysis. JMIR Form Res, 5, e27018. doi:10.2196/27018

Windsor-Shellard, B., & Gunnell, D. (2019) Occupation-specific suicide risk in England: 2011– 2015. The British Journal of Psychiatry, 215, 594–599.

World Health Organization (2021) Suicide worldwide in 2019: global health estimates. Retrieved from https://www.who.int/publications/i/item/9789240026643

Yeh, H.-H., Westphal, J., Hu, Y., et al. (2019) Diagnosed mental health conditions and risk of suicide mortality. Psychiatric Services, 70, 750–757.

Zalsman, G., Hawton, K., Wasserman, D., et al. (2016) Suicide prevention strategies revisited: 10-year systematic review. The Lancet Psychiatry, 3, 646–659. doi:10.1016/S2215-0366(16)30030-X

