## Supplementary File 2 for "Help-seeking needs related to suicide prevention for individuals in contact with mental health services: A rapid scoping review"

| Title |  |
| --- | --- |
| Reviewer Name |  |
| Title |  |
| Year of publication |  |
| Authors |  |
| Country of Origin |  |
| What was the study design? | - Observational - Experimental - Quasi-experimental - Qualitative - Mixed methods or Multi methods - Case study |
| Aim of study |  |
| Study setting |  |
| Sample size |  |
| Reported outcome(s) | Select all   - Suicide - Suicide attempt/behaviour - Suicide ideation |
| Data source | What data did the study collect?   - Health admin - Survey - Interview - Focus group - Other (specify) |
| Service setting |  |
| Population description |  |
| Intersectionality | Based on the description extracted above (population), select all   - Sex - Sexual orientation - Age - Culture - Disability - Education - Ethnicity - Place of residence - Gender - Immigration - Income - Indigeneity - Language - Marital status - Race - Religion - Other: insert (e.g., occupation) |
| Barriers (or risks) to help-seeking needs |  |
| Categorization of barriers | - Individual - Interpersonal - Organization/institutional - Community - Policy/national/law |
| Facilitators to help-seeking needs |  |
| Categorization of facilitators | - Individual - Interpersonal - Organization/institutional - Community - Policy/national/law |
| Implication for help-seeking |  |
| Additional Information about help-seeking |  |
